# Unbiased multi-omics network-based data integration allows clinically relevant outcome-predicting clustering of individuals with heart failure

**DOI:** 10.1101/2025.01.28.25321241

**Authors:** Ekaterina E. Esenkova, Thomas Koeck, Raissa Lerner, Dhanwin Baker, Katrin I. Bauer, Maximilian Nuber, Giorgio Valentini, Laura Bindila, Philipp S. Wild, Elena Casiraghi, Elisa Araldi

**Affiliations:** Preventive Cardiology and Preventive Medicine, Department of Cardiology, University Medical Center of the Johannes Gutenberg University Mainz, Germany; German Center for Cardiovascular Research (DZHK), partner site Rhine-Main, Mainz, Germany; Clinical Epidemiology and Systems Medicine, Center for Thrombosis and Hemostasis (CTH), University Medical Center of the Johannes Gutenberg University Mainz, Germany; Clinical Lipidomics Unit, Institute of Physiological Chemistry, University Medical Center of the Johannes Gutenberg University of Mainz, Mainz, Germany; Department of Computer Science, University of Milan, Milan, Italy; Systems Medicine, Institute of Molecular Biology (IMB), Mainz, Germany; Environmental Genomics and Systems Biology Division, Lawrence Berkeley National Laboratory, Berkeley, CA, USA; Systems Medicine Laboratory, Department of Medicine and Surgery, University of Parma, Parma, Italy

**Keywords:** AI-based omics integration, heart failure, proteomics, lipidomics, clustering, similarity network fusion

## Abstract

Heart failure is a multifaceted clinical syndrome, in which the heart fails to supply adequate blood to meet the body’s oxygen and nutrients needs. Evidence indicates multi-level molecular shifts in heart failure subjects, necessitating unbiased molecular stratification of patients with heart failure. This study utilized AI-based multimodal integration method to analyse 359 lipids and 538 proteins measured in participants of the MyoVasc heart failure cohort. Patient similarity networks were constructed, and spectral clustering, an unsupervised machine learning technique, identified clinically relevant subgroups predictive of patient outcomes. Comparative analyses of cluster-defining proteins and lipids revealed molecular-level insights into heart failure clinical subtypes. In addition to metabolic dysfunctions such as diabetes mellitus, the clinical profiles and outcomes of the identified eight subgroups also showed kidney and liver function indicators. The unbiased molecular characterization was particularly notable in clusters lacking clear, established clinical distinctions, suggesting novel insights into previously uncharacterized patient subgroups. The results show that network-based integration enables to unbiasedly characterize novel molecular subgroups, providing a foundation for improved understanding and management of heart failure.

---

Heart Failure (HF) is a heterogeneous clinical syndrome that is characterized by the inability to pump enough blood (1). The current classification of heart failure is based on the left ventricular ejection faction (LVEF). Patients are subdivided into three categories: HF with preserved ejection fraction (HFpEF) with LVEF ≥ 50%, the mid-range group (HFmrEF) with LVEF 40–49% and reduced ejection fraction (HFrEF) with LVEF < 40% (1). Although the clinical definitions of heart failure phenotypes and stages are well established, there is considerable heterogeneity within these phenotypes in terms of patients’ clinical characteristics and worsening of heart failure prognoses. This heterogeneity results in groups of patients responding differently to treatments and achieving different outcomes. A study by Ahmad et al. demonstrated the superiority of machine learning-based patient clustering over LVEF-based phenotypes in predicting heart failure deterioration (2). The identified clusters were based on clinical and standard blood routine measurements of 44.886 HF patients of the Swedish Heart Failure Registry and were discriminative for survival and response to therapeutics. Using machine learning-based methods integrating other types of data, including molecular data from OMICs platforms which are increasingly rising in quantity and quality, has the potential to stratify patients even more effectively.

Numerous and complex molecular perturbations involving energy metabolism, inflammation and autophagy, among other processes, characterize and precede myocardial dysfunction in heart failure (3–5). Therefore, the inclusion of molecularly defined endotypes could lead to the improvement of existing prevention strategies, including early preventive measures, and contribute significantly to more effective therapies through personalized treatment plans (6). With this in mind, investigating molecular states may be a promising strategy for identifying molecular biomarkers associated with the onset and progression of heart failure.

Previous studies have already attempted to cluster heart failure patients, based on various clinical and biochemical parameters (7–9). Using hierarchical and partitioning clustering methods, researchers have identified clinically relevant subgroups with significant differences in etiology, age, comorbidities, and lifestyle. However, both the sample size and the number of variables were limited in these studies and the focus was only on one of the heart failure phenotypes (e.g. HFpEF, AHF). Another interesting semi-supervised clustering was performed on the nationwide population-based ORISCAV-LUX 2 study (10). The study included 1,356 participants, and with guidance by body mass index (BMI), glycated hemoglobin (HbA1c) and 29 cardiometabolic variables, the authors identified four cardiometabolic clusters. However, the clustering was guided by well-established and limited metabolic parameters and thus, the results can be highly biased.

The current study will overcome some of the limitations of previous clustering attempts by investigating patients in heart failure stages C/D from the MyoVasc cohort (11). MyoVasc is an investigator-initiated, prospective, single-center cohort study based in the city of Mainz, in central-western Germany, featuring sequential deep clinical phenotyping, biobanking, and multi-omics data measurements. It involves all heart failure phenotypes (11), and we apply to it the clinically unbiased approach Similarity Network Fusion (12), which takes advantage of the complementary nature of the OMICs platform to define potentially clinically relevant patient clusters. To confirm the clinical relevance of the computational data-driven analysis, the anamnesis of the subjects, medication intake and risk factors are compared among obtained patient clusters.

## RESULTS

### Data integration and Clustering

The analysis included a subsample of 1,678 subjects from the MyoVasc study (NCT04064450) (11), who were diagnosed with stage C or D heart failure (mean age 67.7 ± 9.9 years; 33.2% women) at baseline and for whom both protein (affinity-qPCR based targeted proteomics) and lipid (4D-LC/TIMS lipidomics (13)) data were available (**Figure 1**).

**Figure 1.**
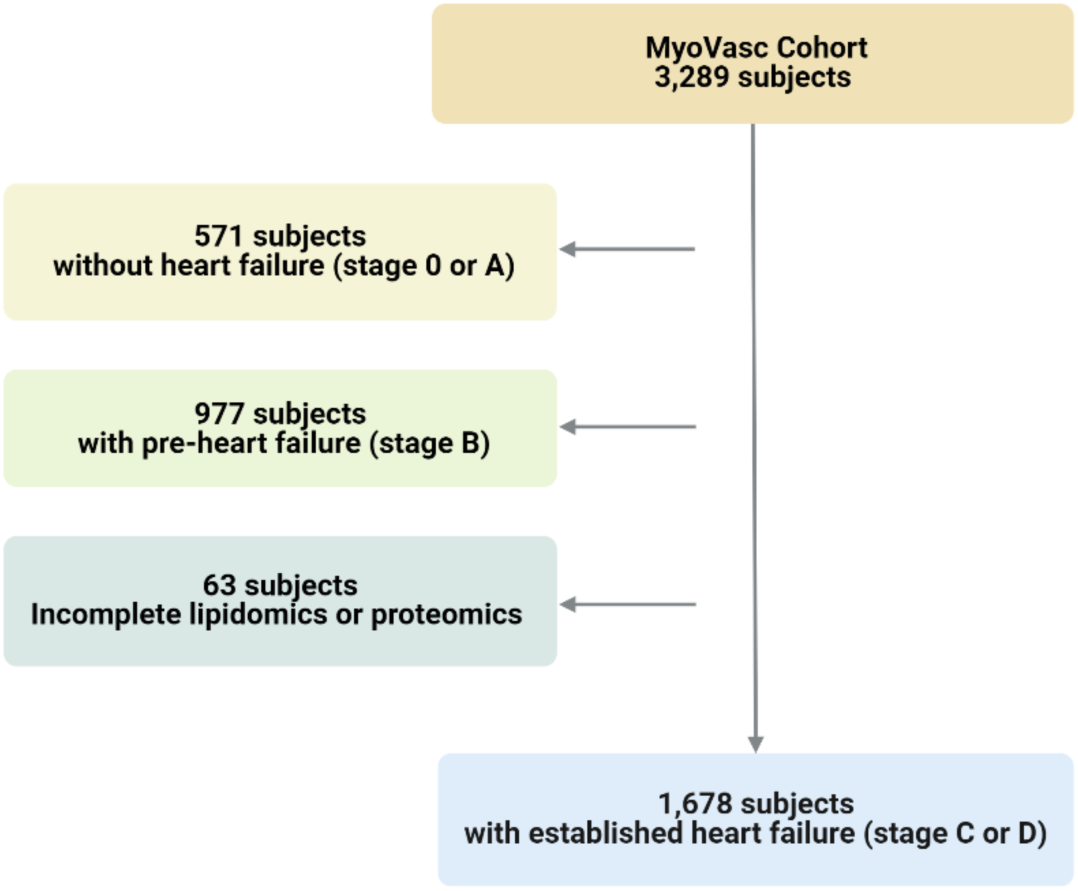
Exclusion criteria and cohort selection. From the entire MyoVasc cohort, we have subsampled 1678 individuals with established heart failure (stage C or D) and complete protein and lipid data.

Each data modality (**Figure 2A**) with 526 proteins and 304 lipids (538 and 359 before pre-processing, respectively), was used to create a separate patient similarity network (PSN), where each node represents a patient and a link between two nodes represents the (protein or lipid) similarity between them (14). The Similarity Network Fusion algorithm (12) was used to integrate the two networks into one - PSN_SNF_ (**Figure 2B**).

**Figure 2.**
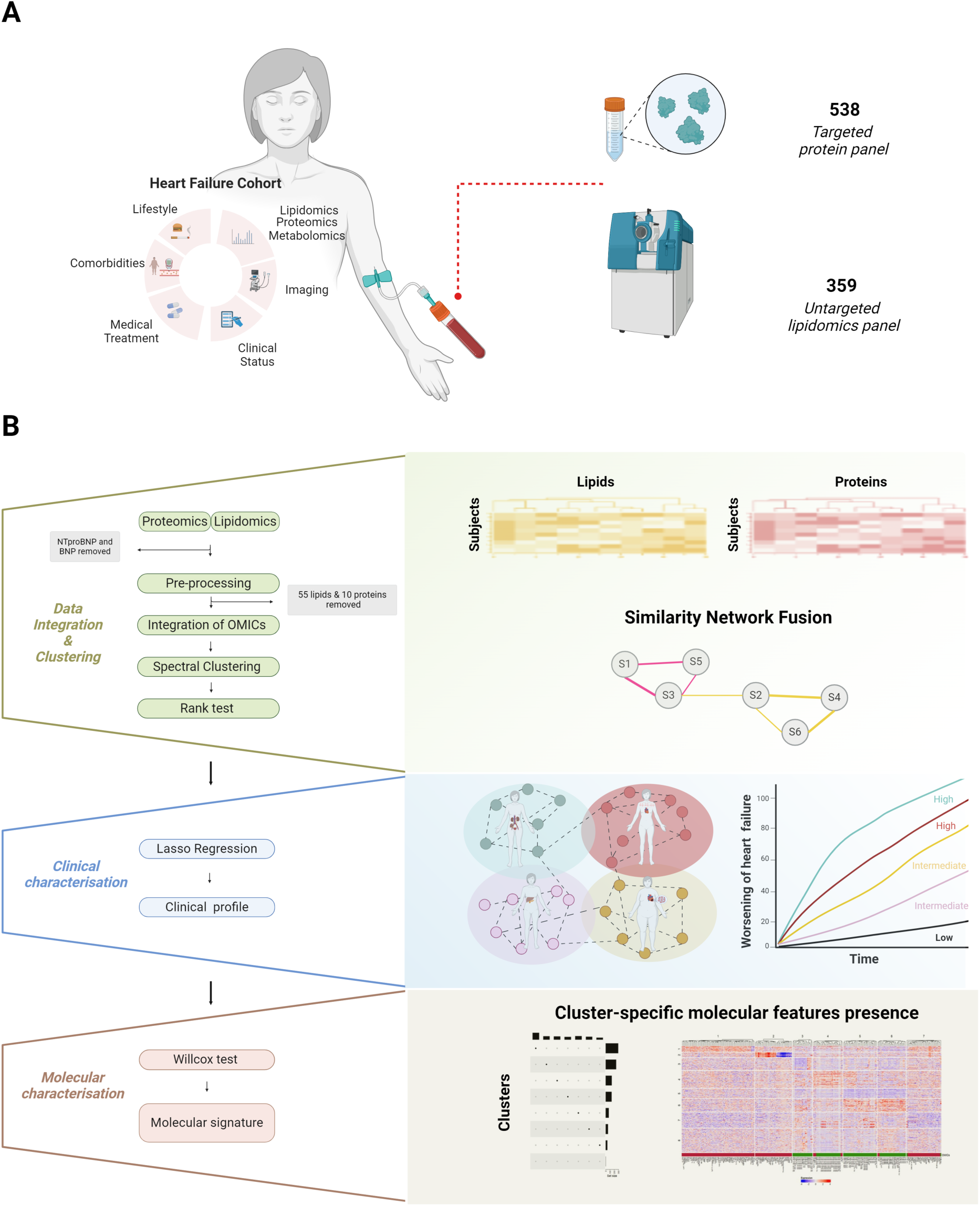
Study design and analysis pipeline. A. MyoVasc subsample of 1678 individuals with 538 proteins and 359 lipids measured. B. The three-parts approach: data integration and clustering, clinical characterization, molecular characterization.

In addition to PSN_SNF_, PSN_avr_ (the average of the protein and lipid PSNs) and PSN_br_ (i.e., the PSN obtained by the standard blood routine measurements) served as methodological and clinical controls, respectively (for more details see Methods section). All three integrated PSNs were then processed by spectral clustering (15), in order to split the data into a number of clusters ranging from 3 to 10. The subjects’ distribution and cluster’s stability were visualized with Sankey plots, color-coded for worsening of heart failure and all-cause death (**Supplementary Figures 1-2**). Due to the complexity of cluster distribution and instability, we have chosen the optimal integration method and number of clusters, based on their ability to differentiate the clinical outcome – worsening of heart failure.

To evaluate the difference in worsening of heart failure between the obtained clusters, we performed a pairwise log-rank test on cumulative incidence curves of worsening of heart failure and results have shown the superiority of OMICs data (both PSN_SNF_ and PSN_avr_) over the blood routine measurements (PSN_br_), demonstrating on average higher mean –log10 of logrank p-values (**Figure 3**). Particularly, the log-rank test showed that the smallest mean p-value is observed with eight clusters integrated with SNF method (**Figure 3**).

**Figure 3.**
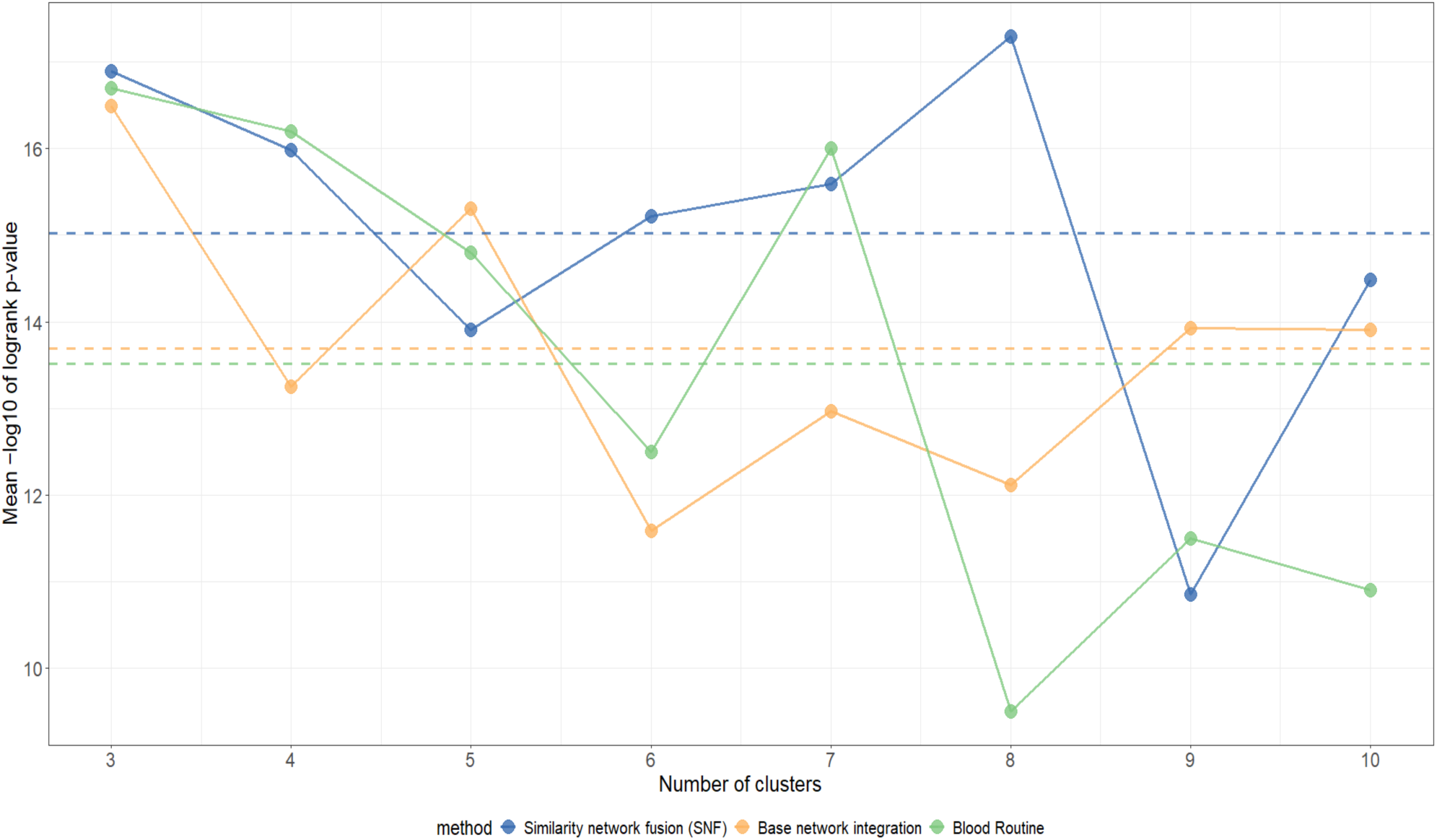
Rank Test. Pairwise log-rank test on cumulative incidence curves of worsening of heart failure was obtained among obtained clusters from integration with Similarity Network Fusion (SNF), Base network integration, and blood routine, and mean –log10 of logrank p-values was plotted. A higher mean –log10 of logrank p-value defines better separation of clusters for the worsening of heart failure outcome (4 year follow-up). With best result being the application of Similarity Network Fusion (SNF) to integrate the omics data, followed by dividing it into 8 clusters.

This demonstrated that eight clusters were sufficient for optimal clinical separation of subjects, based on worsening of heart failure (**Figure 4A**). Cluster 8, which was characterized by the lowest incidence of worsening of heart failure outcomes and an even distribution of women and men, was selected as the reference cluster for further clinical characterization (**Supplementary Table 1**). In addition, multivariable Cox proportional hazard regression analyses (adjusted for age, sex and NT-proBNP*) demonstrated that the first four clusters are significantly different for worsening of heart failure in comparison to the reference cluster (**Figure 4B**). NT-proBNP* stands for residuals of linear model function, where log transformed NT-pro-BNP values are dependent variables on estimated glomerular filtration rate (eGFR). It is important to note that the patients in these clusters were not significantly older than in the reference cluster 8. On the contrary, the mean age value in cluster 4 was significantly lower than in cluster 8 (**Figure 4C**). Further, we tried to investigate the possible clinical and molecular rationale for the differences in worsening of heart failure of the four clusters with worse prognosis.

**Figure 4.**
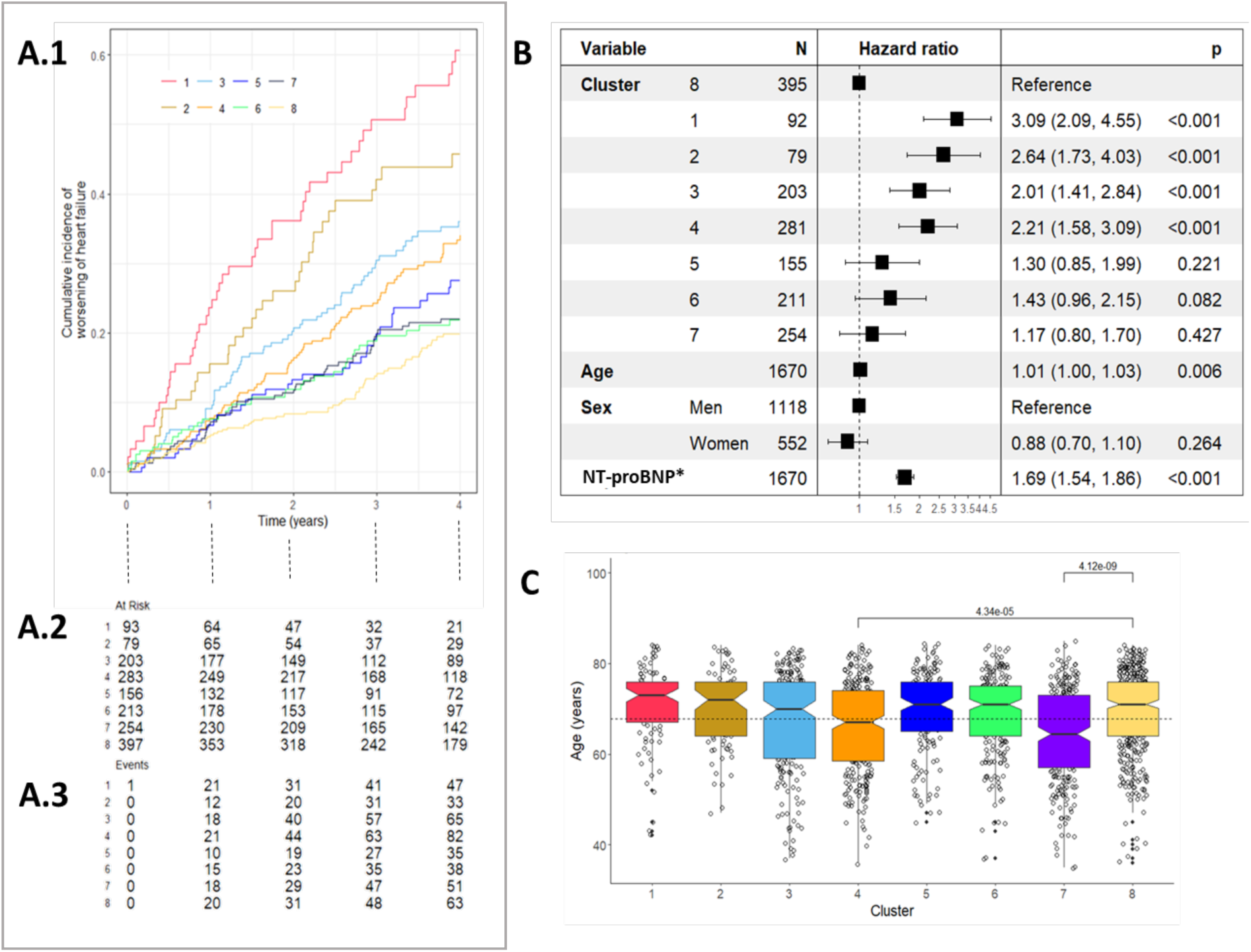
Clinical characteristics of clusters, based on cumulative incidence of worsening of heart failure. A.1 Cumulative incidence of worsening of heart failure (4 years follow-up) for 8 clusters subdivision. A.2 Number of subjects at risk of worsening of heart failure for 4 years. A.3 Number of subjects with worsening of heart failure for 4 years. B. Forest plot of Cox regression, where NT-pro-BNP is defined as residuals of linear model function, where log transformed NT-pro-BNP values are dependent variable on estimated estimated glomerular filtration rate (eGFR). C. Boxplot of age distribution for each cluster. The mean age of Cluster 4 and cluster 7 is significantly lower than cluster 8 (reference).

### Clinical characteristics of clusters with worse predicted outcome

Multinomial logistic Lasso regression defined the discriminating clinical characteristics for each cluster (**Figure 5**), indicating that each cluster represents a combination of different comorbidities, which potentially contribute to the progression of heart failure. Namely, cluster 1 is enriched in subjects with chronic kidney disease (CKD), venous thromboembolism (VTE), atrial fibrillation (AF) and fatty liver. Impaired renal function was further confirmed by elevated urea and creatinine levels, and elevated Gamma-Glutamyl Transferase (GGT) pointed towards liver dysfunction (**Supplementary Table 2**). In addition, we observed significantly elevated levels of C-reactive protein (CRP). Cluster 2 was also characterized by the presence of chronic kidney disease and atrial fibrillation, but additionally included subjects with a history of stroke. Cluster 3 consists of subjects with obesity and history of myocardial infarction. However, unlike Cluster 4, this group does not show severe metabolic disturbances, such as elevated HbA1c or HOMA-IR levels (**Supplementary Table 2**). The comorbidities in cluster 4, including fatty liver, dyslipidemia, diabetes mellitus, and arterial hypertension, indicate metabolic syndrome as a characteristic feature. Peripheral arterial disease and smoking as a lifestyle factor were also present. The clinical parameters of cluster 4 including elevated HbA1C, HOMA-IR,

**Figure 5.**
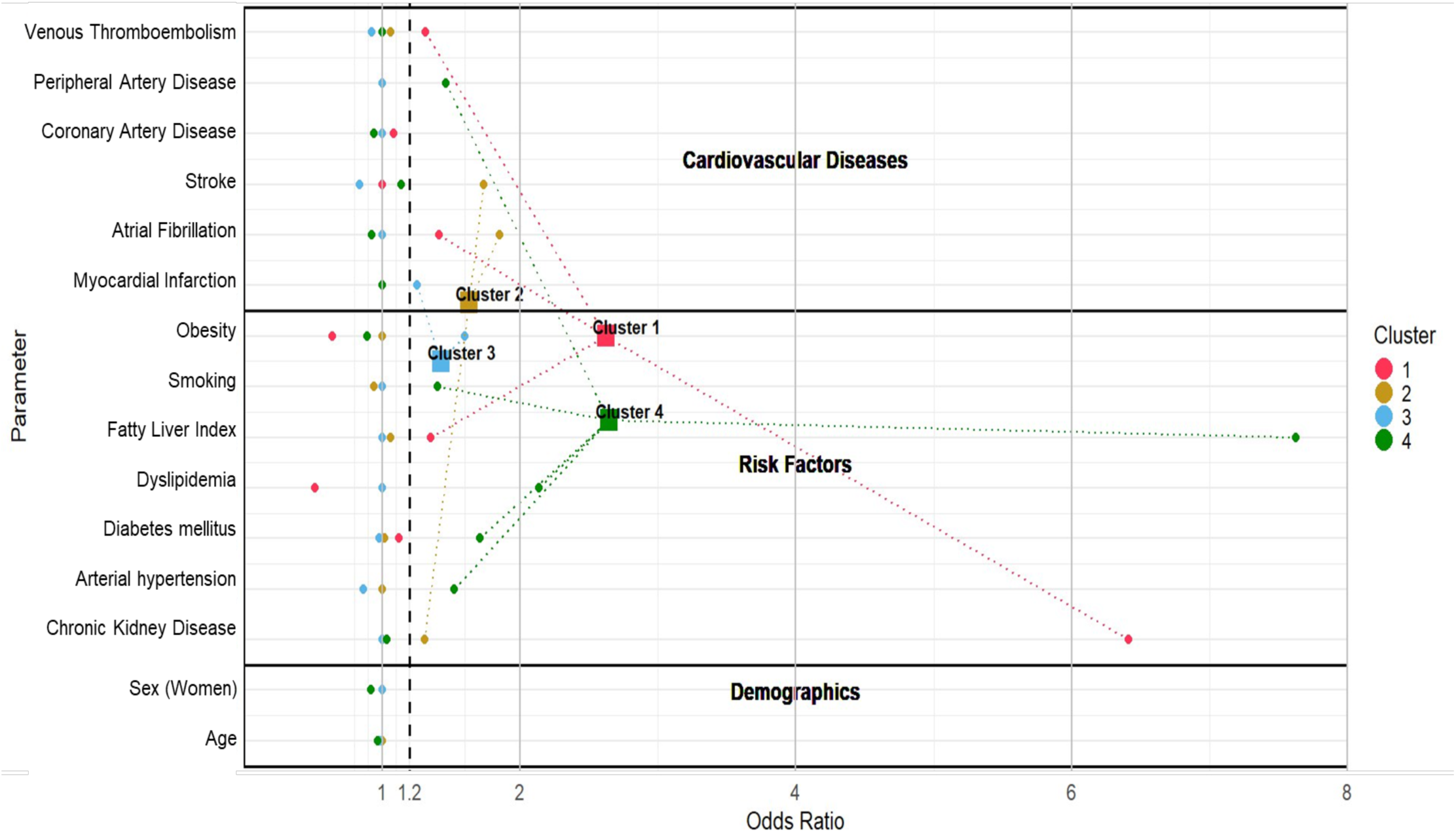
Clinical characteristics of clusters. The odds ratio values are based on multinomial logistic lasso regression. The cluster label boxes are calculated as centroids to the significant parameters (that have odds ratio C-peptide and triglycerides, confirmed the metabolic syndrome as the main characteristic feature (**Supplementary Table 2**).

### Molecular characteristics of clusters with worse predicted outcome

Data integration and clustering were based solely on molecular features. We performed a paired Wilcoxon test to identify specific molecular features discriminating each cluster. The fused network approach captures both shared and complementary information from different OMICs platforms, so we anticipated that a combination of protein and lipid features would define each cluster. Interestingly, the two clusters with the highest incidence of worsening heart failure (clusters 1 and 2) were characterized exclusively by protein features — 69 and 35 proteins, respectively (**Figure 6**). The majority of these proteins are associated with inflammatory processes. Particularly, most proteins in cluster 1 belong to the interleukin family, while those in cluster 2 are part of the tumor necrosis factor (TNF) superfamily (**Figure 6, Supplementary Figures 3-4**).

**Figure 6.**
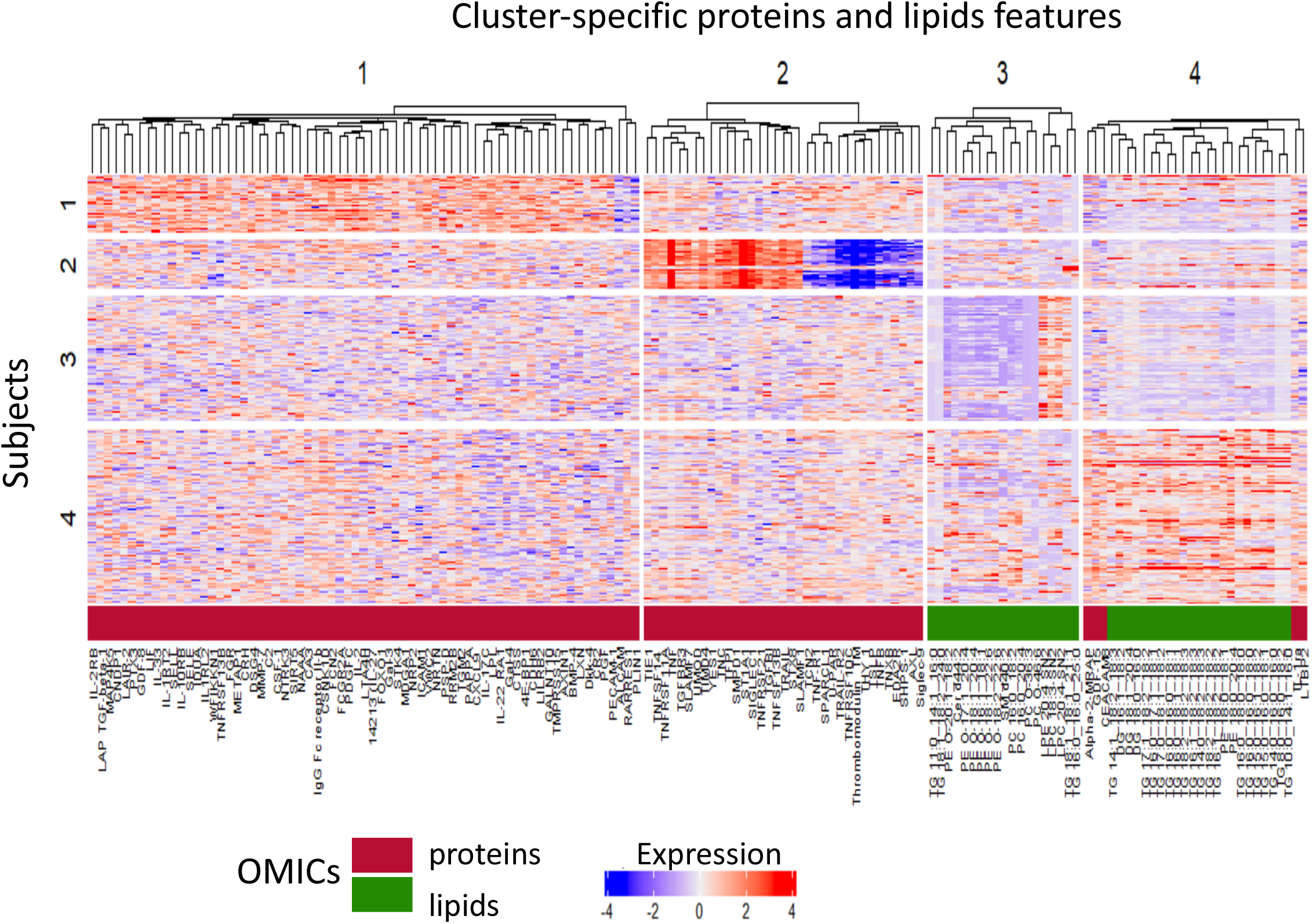
Cluster-specific proteins and lipids signatures. In the post-hoc analysis, unique protein and lipid sets for each cluster were identified using pairwise Wilcoxon tests, with p-values adjusted by the Bonferroni-Holm correction. The integration and clustering allow the mixing of the two platforms, however, only cluster 4 demonstrated a combined profile of lipids and proteins.

In contrast, the molecular signature of cluster 3 comprises only 19 lipid species (**Supplementary Figure 5, Supplementary Table 4**), including lysophosphatidylcholine (LPC), phosphatidylcholine (PC), sphingomyelin (SM), phosphatidylethanolamine (PE), triglycerides (TG), ceramides (Cer), and lysophosphatidylethanolamine (LPE). Aligned with the complementary nature of the Similarity Network Fusion (SNF) method, the molecular profile of cluster 4 combines 5 proteins and 23 lipids, indicating a potential interplay between lipids and proteins at the enzymatic level and beyond (**Supplementary Figure 6, Supplementary Table 4**). Moreover, cluster 4 features a significant enrichment of Growth Differentiation Factor 15 (GDF-15) (**Supplementary Figure 6A**), even adjusted for metformin intake (**Supplementary Figure 7**)

## DISCUSSION

In this study, we applied an unbiased integration approach, Similarity Network Fusion (SNF) (**Figure 2B**) that is distinct and superior to simple integration techniques due to several reasons. First, it uses networks as a basis for integration and thus, it is a robust method to the limited sample size, noise and data heterogeneity that are present in cohorts with clinical syndromes. Secondly, the parallel interchanging diffusion processes that simultaneously update the protein and lipid matrices allows to identify not yet experimentally proven connections between lipid and protein spaces. The availability of multi-omics data, combined with detailed clinical profiles, is a significant advantage of the MyoVasc cohort. As the results have demonstrated, the richer variable space (including untargeted features) of the multi-omics approach outperforms the limited set of clinically established blood routine measurements in regards to the clinical separation of clusters, based on worsening of heart failure (log-rank test on clusters worsening of heart failure incidence) (**Figure 3**). Although the clinical profiles of worse-performing clusters confirm the previously established clinical comorbidities of heart failure, there are also clusters with worse outcomes and a lack of defined heart failure risk factors (**Figure 5**). Such patients’ profiles are challenging to discern at the doctor’s visit alone and benefit the most from the in-depth molecular characterization. With this in mind, the molecular characterization and its bridging to the clinical profile is a crucial part of the analysis.

The majority of upregulated proteins in Cluster 1 belong to the Interleukin family, which are involved in inflammatory and immune responses (**Figure 6**, **Supplementary Figure 3**). Consistent with this, C-reactive protein (CRP) levels are elevated in this cluster (**Supplementary Table 2, Supplementary Figure 3**). The remaining proteins are implicated in broader physiological processes, including cell growth, differentiation, signaling, and fibrosis. Proteins with more specific relevance to renal function include Carbonic Anhydrase 3 (CA3), which is essential for bicarbonate transport and maintenance of acid-base balance (16), and Lipoprotein Lipase (LPL), which is involved in lipid metabolism (17). Interestingly, among three down-regulated proteins, we were able to identify Perilipin-1 (PLIN1). It is a regulator of adipocyte lipid metabolism, when down-regulated, can result in uncontrolled lipolysis (18), which, particularly in diabetic nephropathy, has been associated with increased lipid accumulation and inflammation in renal tissues (19).

As Cluster 2 is clinically characterized by the prevalence of atrial fibrillation (AF) and a history of stroke, it is expected for the protein panel to reflect the related processes, such as structural and electrical remodeling of the heart, inflammation and fibrosis. The majority of the differential proteins in cluster 2 are part of the tumor necrosis factor (TNF) superfamily (**Supplementary Figure 4**). Specifically, these include cytokines and ligands such as TNF (Tumor Necrosis Factor), LTA (Lymphotoxin-alpha), TNFSF13B (BAFF), TNFSF14 (LIGHT), and TNFSF10 (TRAIL), as well as several TNF receptors. These receptors include TNFRSF10B (DR5) and TNFRSF10A (DR4), which initiate apoptosis; TNFRSF10C (DcR1), which inhibits apoptosis; and TNFRSF11A (RANK) and TNFRSF1A, which mediate various effects like inflammation and proliferation. The upregulation of those proteins indicates chronic inflammation and potential loss of cardiomyocytes, triggering atrial fibrillation (20), which is the top clinical parameter in cluster 2. Additionally, there is a connected network, involving PLAUR (Plasminogen Activator, Urokinase Receptor), THBD (Thrombomodulin), and TFPI (Tissue Factor Pathway Inhibitor) (**Supplementary Figure 4**). These proteins regulate blood coagulation and fibrinolysis, with their dysregulation potentially leading to thromboembolic events such as stroke (the second top clinical feature in cluster 2). The inflammatory environment that promotes AF development can be linked to increased levels of immune-modulatory proteins such as SLAMF1 (Signaling Lymphocytic Activation Molecule Family Member 1) and SLAMF2 (CD48).

Cluster 3 is characterized by a higher prevalence of obesity and myocardial infarction (MI) among subjects. However, unlike Cluster 4, this group does not show severe metabolic disturbances, such as elevated HbA1c or HOMA-IR levels (**Supplementary Table 2**). Molecular analysis identified 19 lipid species within this cluster (**Supplementary Figure 5**), including lysophosphatidylcholines (LPC), phosphatidylcholines (PC), sphingomyelins (SM), phosphatidylethanolamines (PE), triglycerides (TG), ceramides (Cer), and lysophosphatidylethanolamines (LPE). The analysis has shown that their structure comprises 4 saturated (undecanoic acid (FA 11:0), palmitic acid (FA 16:0), lignoceric acid (FA 24:0), and pentacosanoic acid (FA 25:0)), 6 unsaturated (myristoleic acid (FA 14:1), palmitoleic acid (FA 16:1), heptadecenoic acid (FA 17:1), oleic acid (FA 18:1), gondoic acid (FA 20:1), and heneicosenoic acid (FA 21:1)) and 4 polyunsaturated acids (Alpha-linolenic acid (FA 18:3), arachidonic acid (FA 20:4), linoleic acid (FA 18:2), and docosahexaenoic acid (FA 22:6) (**Supplementary Table 4**). Strikingly, out of 14 unique fatty acids that are part of these lipids structure, 3 are very long-chain fatty acids (VLCFAs), comprising docosahexaenoic (DHA; FA 22:6), tetrasanoic (FA 24:0) and pentacosanoic acid (FA 25:0). Interestingly, docosahexaenoic fatty acid (DHA) is present as a part of phosphatidylethanolamines (PE) that are reduced in cluster 3. This finding partly corresponds to the results of Alter et al., where overall levels of DHA in the lipidome were decreased in patients with heart failure (21). Although VLCFAs are gaining evidence to be associated with lower risk of incident HF and diabetes mellitus in previous studies (22), their health benefits are yet to be investigated, particularly in relation to their saturation degree.

In comparison to cluster 3, cluster 4 is characterized by severe disturbances in its metabolic parameters (**Supplementary Table 2**). The majority of molecular features are lipids and can be subdivided into 3 functional subgroups: two phosphatidylethanolamines, three diglycerides and 18 triglycerides (**Supplementary Figure 6, Supplementary Table 4**). They contain 7 saturated fatty acyls: (caprylic acid (FA 8:0), decanoic acid (FA 10:0), myristic acid (FA 14:0), pentadecanoic acid (FA 15:0), palmitic acid (FA 16:0), heptadecanoic acid (FA 17:0), stearic acid (FA 18:0)), 4 unsaturated fatty acyls: (myristoleic acid (FA 14:1), palmitoleic acid (FA 16:1), oleic acid (FA 18:1)), and 3 polyunsaturated fatty acyls: (linoleic acid (18:2), alpha-linolenic acid (18:3), and arachidonic acid (20:4)). Related to chain length, lipid containing short-chain fatty acids (SCFAs) were not detected under these experimental conditions, whereas lipids containing 2 medium-chain fatty acids (MCFAs) and 12 long-chain fatty acids (LCFAs) (Supplementary Table 4) were detected. The prevalence of long-chain fatty acids (LCFAs) has been shown to promote lipid accumulation and insulin resistance in comparison to medium-chain fatty acids (MCFAs), which are more efficiently absorbed (23).

In addition to 23 lipids, cluster 4 is further characterized by five up-regulated proteins: GDF-15, Alpha-2-MRAP, CEACAM8, LTBP2, IL-1RA (**Supplementary Figure 6**). The most expressed protein GDF-15 has already been proposed as a potential biomarker for heart failure, obesity, and insulin resistance (24). Growth differentiation factor 15 (GDF15) is a cytokine that is believed to be involved in food control and its expression can be induced by cellular and metabolic stress. Apart from the disease state, its significant upregulation in cluster 4 can be also explained by the evidence that GDF-15 is produced by the kidney, as a response to metformin treatment. Cluster 4 has the highest proportion of metformin users, with 31 out of 82 individuals taking metformin belonging to this cluster. However, the residuals from the regression of GDF-15 expression on metformin intake (binary variable) still demonstrated the significant increase of GDF-15 in cluster 4 (**Supplementary Figure 7**). The upregulation of the Interleukin-1 receptor antagonist is not surprising as a response to central inflammation, potentially triggered by IL1 (which is not measured due to instability of protein). The Danish clinical trial has demonstrated the beneficial role of IL1-RA supplementation (anakinra) in type 2 diabetic patients via blockage of interleukin-1 (25). The contribution of the other proteins to heart failure or metabolic regulation is less direct. Alpha-2-MRAP is a molecular chaperone that assists the folding and trafficking of LDL receptor-related proteins (26). Unlike CEACAM1, which is involved in insulin regulation (27), the role of CEACAM8 in metabolic disorder and heart failure is not yet established. It is expressed on activated neutrophils, and thus, can contribute indirectly via inflammation processes. CEACAM8 is an intriguing protein for further investigation. The Homage study identified it as one of 38 proteins contributing to the proteomic profile associated with incident heart failure in the replicated analysis phase (28). LTBP2 is expressed in elastic tissues, such as the heart and is involved in cardiac tissue fibrosis and remodeling - hallmarks of heart failure (29, 30).

To conclude, this study leverages the large number of variables from a deeply clinically and molecularly characterized heart failure cohort, the MyoVasc study, together with the power of molecular data integration by Similarity Network Fusion (SNF) to identify novel heart failure patient subtypes and discriminating protein and lipid biomarkers that can be used in the clinic to tailor treatments and predict outcomes. We have demonstrated that Similarity Network Fusion (SNF) enables more accurate patient stratification by integrating multiple types of OMICs data, and further analyses integrating additional molecular datasets will enhance the identification of distinct molecular subgroups of heart failure patients for personalized treatment approaches.

## Methods

### Study cohort

Data from the MyoVasc study (NCT04064450), a prospective cohort study of heart failure (HF) was analyzed (11). During a five-hour baseline examination in the dedicated study center in Mainz (central-western Germany), each participant underwent a deep clinical phenotyping, including plasma samples collection (stored in the Biobank for further molecular characterization).

### Proteomics

Once-thawed EDTA-anticoagulated blood plasma samples were analyzed with proximity extension assay technology (Olink Biosciences, Uppsala, Sweden), a targeted method of protein expression quantification that produces normalized expression values (31). In total six panels (trade names: Cardiovascular II, Cardiovascular III, Cardiometabolic, Inflammation, Immune Response, Organ Damage) were measured and used for these analyses.

### Lipidomics

Lipid extraction from blood plasma was performed using the automated high-throughput extraction platform described by Lerner et al. (13). Lipid analysis was conducted using 4D-LC-TIMS lipidomics. The samples were analyzed using Elute UHPLC-TIMS-TOF Pro instrument (Bruker Daltonics, Germany) operated in both negative and positive ionization modes. Instrumental parameters for data acquisition and lipid data processing followed the protocol described by Lerner et al. (13). Lipid features were identified and annotated based on mass accuracy, RT, CCS, and characteristic fragment ions observed in their MS² spectra obtained by PASEF. The lipid classes ceramides (Cer), glycerophosphoethanolamines (PE), and glycerophosphoinositols (PI) were analyzed and quantified in negative ion mode, while glycerophosphocholines (PC), sphingomyelins (SM), cholesterol, triacylglycerols (TG), and diacylglycerols (DG) were analyzed and quantified in positive ion mode. Lipid quantification was carried out using level-2 deuterated internal standards, with the exception of cholesterol, for which the level-1 internal standard was used, and DG, for which a level-3 internal standard was applied. A multi-point calibration strategy using external lipid standards was applied to all classes, except for CE, DG, TG, and Cer, where a single-point calibration was performed.

### Data handling and statistical analysis

The MyoVasc cohort was subsampled to the individuals with C/D stage heart failure at the baseline visit. Prior to the data integration step, the proteomics and lipidomics data were preprocessed. Namely, it included the following steps: data imputation by using missForest (32), chosen due to its superior robustness in several medical applications (33, 34); identification and removal of either highly correlated predictors (pairwise correlation cutoff >= 0.9), which would cause inflated and/or biased estimates, or not-informative predictors, i.e. predictors being practically constant (with near zero variance) (15).

After pre-processing, the established gold standards of heart failure NT-proBNP and BNP were removed from the protein matrix to avoid any clinical bias in the following analyses. To obtain an integrated PSN, capturing both local and global pairwise similarities, Similarity Network Fusion (SNF, see subsequent Section “Similarity Network Fusion”) was used, due to its effectiveness in several molecular and multi-omics applications (12). To assess and validate the usage of SNF, the results obtained on the SNF-integrated PSN, hereafter referred to as PSN_SNF_, were compared to those obtained by computing individual protein and lipids PSNs (affinity matrices) - by using the same scaled Euclidean kernel used by SNF (12), and then integrating them with basic network integration techniques (the average of the protein and lipids PSNs - PSN_avg_). As a clinical control, we also created a PSN using the standard blood routine measurements (combination of cell count and humoral biomarkers, excluding NT-pro-BNP and troponin) - PSN_br_ (Supplementary Table 2, Table 3).

Spectral Clustering was then used to cluster the integrated affinity matrices PSN_SNF_, PSN_avg_ and PSN_br_ (12). The choice of the optimal integration method and of the number of clusters for the further clinical analysis was based on the smallest log-rank test p-value in the outcome worsening of heart failure. It evaluated the average significance of the difference in worsening of heart failure profiles from three to ten clusters division. For the chosen method and number of cluster we first calculated the incidence of worsening of heart failure and then performed the multivariable Cox proportional hazard regression model of each cluster against the reference cluster (35). The reference cluster was defined as a cluster with the lowest mean cumulative incidence of worsening of heart failure. The Cox regression was adjusted to age, sex and NT-pro-BNP*. NT-pro-BNP* was defined as residuals of linear model function, where log transformed NT-pro-BNP values are dependent variables on estimated glomerular filtration rate (eGFR).

### Similarity Network Fusion (SNF)

SNF integrates multiple data sources through a cross-diffusion process that iteratively exchanges information among the Patient Similarity Networks (PSNs) constructed from each data source (12).

More specifically, given *m* data sources, SNF first applies a scaled exponential affinity kernel to construct an individual (unimodal) PSN, represented as *W*^(*s*)^, *s* ∈ {1, … , *m*}, for each data source *s*. These PSNs are subsequently processed to generate:

1. *P*^(*s*)^, a normalized PSN capturing ‘’global’’ relationships among patients; and
2. *S*^(*s*)^, a ‘’local’’ PSN representing topologies within neighborhoods in *s*.

For two points *x_i_* and *x_k_*, *S*^(*s*)^(*i*, *k*) > 0 holds only if *x_k_* ∈ *N_i_* , where *N_i_* is the k-nearest neighborhood of *x_i_* in source *s*.

In the simplest case of two sources *s* ≠ *v*, the t-th iteration of the diffusion process updates *P_t_*^(^*^s^*^)^ and *P_t_*^(^*^t^*^)^ using the following equations:

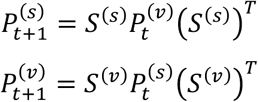

where *X^T^* is the transpose of matrix *X* Focusing on source *s* and two points *x_i_* and *x_j_*, the following equation can be expanded for a single element *P_t_*^(^*^s^*^)^(*i*, *j*) as:

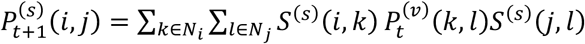

Here, the product inside the summations is non-zero only when *x_k_* ∈ *N_i_* and *x_l_*, ∈ *N_j_* are also global neighbors in source *v* during the previous iteration (*P_t_*^(*v*)^(*k*, *l*) ≠ 0). In essence, similarity information is exchanged between sources *v* and *s* only through their shared global and local neighborhoods. This iterative process progressively aligns global similarities across sources. At convergence — or after *T* iterations—the integrated (consensus) PSN, *P*^(*c*)^, is calculated as the average of *P*^(*s*)^ and *P*^(*v*)^; in other words, i.e., in the case of *m* = 2 sources, *s* and *v*:

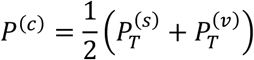

*P*^(*c*)^ captures shared and complementary global and local information from all individual data sources.

### Clinical and molecular characterization

To identify clinical features characterizing each cluster, we represented each individual by using the main comorbidities that contribute to the heart failure onset and development and performed multinomial logistic Lasso regression between each cluster and the reference cluster, that has shown the lowest incidence of worsening of heart failure, no significant age difference and an even distribution of women and men (36). To confirm the clinical assignment or/and to discover new distinguishing phenotypical features of the clusters, the variables from blood routine and cardiological assessment were also compared.

After clinical characterization, we investigated the molecular profiles of each cluster. Performing post-hoc-analysis by applying pairwise Wilcoxon tests with p value adjusted by Bonferroni-Holm correction, we obtained the set of molecular features (proteins, lipids) characterizing each cluster. In the first selection round, we have kept the features that would show significant differences to all seven clusters per comparison, and then we focused on the ones that show this uniquely for one cluster. For each cluster, we defined a list of such features, and the further biological analysis was done, using STRING database to create protein-protein interaction networks for proteins with a minimum required interaction score of 0.4 (37) and literature search to characterize different lipid classes. Additionally, the lipid classes space was transferred into fatty acids space: each lipid formula was subdivided into corresponding fatty acyls and each fatty acyl was further classified for saturation and chain length. Chain length classes were defined based on the number of carbons as follows: short-chain fatty acids (SCFAs) with 1 to 6 carbons (C1–C6), medium-chain fatty acids (MCFAs) with 7 to 12 carbons (C7–C12), long-chain fatty acids (LCFAs) with 13 to 21 carbons (C13–C21), and very long-chain fatty acids (VLCFAs) with 22 to 28 carbons (C22–C28).

## Data Availability

This project constitutes a major scientific effort with high methodological standards and detailed guidelines for analysis and publication. Data are not made available for the scientific community outside the established and controlled workflows and algorithms. To meet the general idea of verification and reproducibility of scientific findings, we offer access to data at the local database in accordance with the ethics vote on request (contact: info@myovasc).

## Code availability

Analyses were conducted using R 4.2.1. The dataset and codes, containing de-identified data, and the R file used to generate the results in this paper are both available upon reasonable request and sent to the corresponding author.

## Acknowledgments

The authors express their gratitude to the study participants and the current as well as former members of the MyoVasc study team. Part of this work is included in the thesis of Ekaterina Esenkova. We acknowledge the technical help of Claudia Schwitter for extraction and of lipids from MyoVasc cohort.

## Authors’ contributions

EEE conceived and designed the study and the analyses, performed the analyses and interpreted the data, prepared the figures, and wrote the manuscript. TK optimized protocols, prepared the samples for OLINK proteomics, contributed to OLINK data generation, pre- and post-processing. DB and RL prepared the samples for 4D-TIMS Lipidomics, contributed to 4D-TIMS Lipidomics data generation, pre- and post-processing, and analyzed data. LB acquired funding, conceived and designed the study, supervised the acquisition and analyses of 4D-TIMS Lipidomics data, and interpreted the data. PSW acquired funding, conceived and designed the study, supervised the analyses and interpreted the data. EC supervised the analysis and interpreted the data. EA acquired funding, conceived and designed the study and the analyses, supervised the statistical analyses, interpreted the data, and wrote the manuscript. All authors read and approved the final manuscript.

## Ethical approval consent to participate

The local data protection officer and the responsible ethics committee approved the study protocol [ref. no. 2018-13064] prior to study initiation. All study participants provided written informed consent prior to study enrolment. The Declaration of Helsinki^38^ and the recommendations of good clinical practice and good epidemiological practice were followed in all study procedures.

## Rights and permissions

**Open Access** This article is licensed under a Creative Commons Attribution 4.0 International License, which permits use, sharing, adaptation, distribution and reproduction in any medium or format, as long as you give appropriate credit to the original author(s) and the source, provide a link to the Creative Commons licence, and indicate if changes were made. The images or other third party material in this article are included in the article’s Creative Commons licence, unless indicated otherwise in a credit line to the material. If material is not included in the article’s Creative Commons licence and your intended use is not permitted by statutory regulation or exceeds the permitted use, you will need to obtain permission directly from the copyright holder. To view a copy of this licence, visit http://creativecommons.org/licenses/by/4.0/.

## SUPPLEMENTARY TABLES

**Supplementary Table 1.** Baseline characteristics of clustering-based subgroups (categorical variables)

| Characteristics | Analysis sample | Cluster 1 | Cluster 2 | Cluster 3 | Cluster 4 | Cluster 5 | Cluster 6 | Cluster 7 | Cluster 8 |
| --- | --- | --- | --- | --- | --- | --- | --- | --- | --- |
| *n* | 1,678 | 93 | 79 | 203 | 283 | 156 | 213 | 254 | 397 |
| *Demographics* |  |  |  |  |  |  |  |  |  |
| Age [years] mean±SD | 67.7±9.9 | 70.8±8.7 | 70.0±8.6 | 67.3±11.3 | 65.9±9.9 | 69.5±9.0 | 68.7±9.0 | 64.2±10.4 | 69.2±9.2 |
| Sex (Women) | 33.2 (557) | 30.1 (28) | 31.6 (25) | 30.0 (61) | 21.6 (61) | 9.0 (14) | 67.6 (144) | 16.9 (43) | 45.6 (181) |
| *Cardiovascular risk factors, % (n)* |  |  |  |  |  |  |  |  |  |
| Arterial hypertension | 82.5 (1385) | 84.9 (79) | 82.3 (65) | 79.3 (161) | 90.5 (256) | 84.6 (132) | 82.2 (175) | 78.0 (198) | 80.4 (319) |
| Diabetes mellitus | 29.7 (499) | 38.7 (36) | 36.7 (29) | 28.6 (58) | 51.2 (145) | 37.2 (58) | 16.9 (36) | 22.8 (58) | 19.9 (79) |
| Dyslipidaemia | 79.9 (1341) | 68.8 (64) | 79.7 (63) | 82.3 (167) | 93.6 (265) | 90.4 (141) | 72.3 (154) | 83.5 (212) | 69.3 (275) |
| Obesity | 38.5 (646) | 28.0 (26) | 36.7 (29) | 46.3 (94) | 52.7 (149) | 42.9 (67) | 34.3 (73) | 33.1 (84) | 31.2 (124) |
| Smoking | 13.8 (232) | 14.0 (13) | 8.9 (7) | 14.8 (30) | 20.2 (57) | 16.0 (25) | 6.6 (14) | 19.3 (49) | 9.3 (37) |
| *Medical history, % (n)* |  |  |  |  |  |  |  |  |  |
| Cardiovascular disease | 94.5 (1586) | 94.6 (88) | 98.7 (78) | 96.1 (195) | 95.1 (269) | 98.7 (154) | 89.2 (190) | 95.3 (242) | 93.2 (370) |
| Atrial fibrillation | 33.6 (564) | 51.6 (48) | 53.2 (42) | 32.0 (65) | 30.4 (86) | 33.3 (52) | 32.4 (69) | 28.0 (71) | 33.0 (131) |
| Congestive heart failure | 85.8 (1439) | 93.5 (87) | 89.9 (71) | 82.3 (167) | 87.3 (247) | 91.0 (142) | 82.2 (175) | 81.5 (207) | 86.4 (343) |
| Coronary artery disease | 52.4 (879) | 55.9 (52) | 50.6 (40) | 57.1 (116) | 58.7 (166) | 69.2 (108) | 32.9 (70) | 60.2 (153) | 43.8 (174) |
| History of MI | 35.4 (594) | 35.5 (33) | 32.9 (26) | 42.9 (87) | 38.9 (110) | 49.4 (77) | 22.1 (47) | 41.7 (106) | 27.2 (108) |
| History of stroke | 11.3 (190) | 16.1 (15) | 22.8 (18) | 7.9 (16) | 12.0 (34) | 10.3 (16) | 14.6 (31) | 9.4 (24) | 9.1 (36) |
| Peripheral artery disease | 10.4 (174) | 16.1 (15) | 11.4 (9) | 11.3 (23) | 17.3 (49) | 11.5 (18) | 9.4 (20) | 7.1 (18) | 5.5 (22) |
| Chronic Kidney Disease | 22.3 (375) | 68.8 (64) | 32.9 (26) | 21.7 (44) | 26.5 (75) | 24.4 (38) | 16.4 (35) | 9.4 (24) | 17.4 (69) |
| History of cancer | 18.7 (314) | 28.0 (26) | 21.5 (17) | 16.3 (33) | 19.1 (54) | 16.7 (26) | 20.7 (44) | 13.8 (35) | 19.9 (79) |

**Supplementary Table 2.**
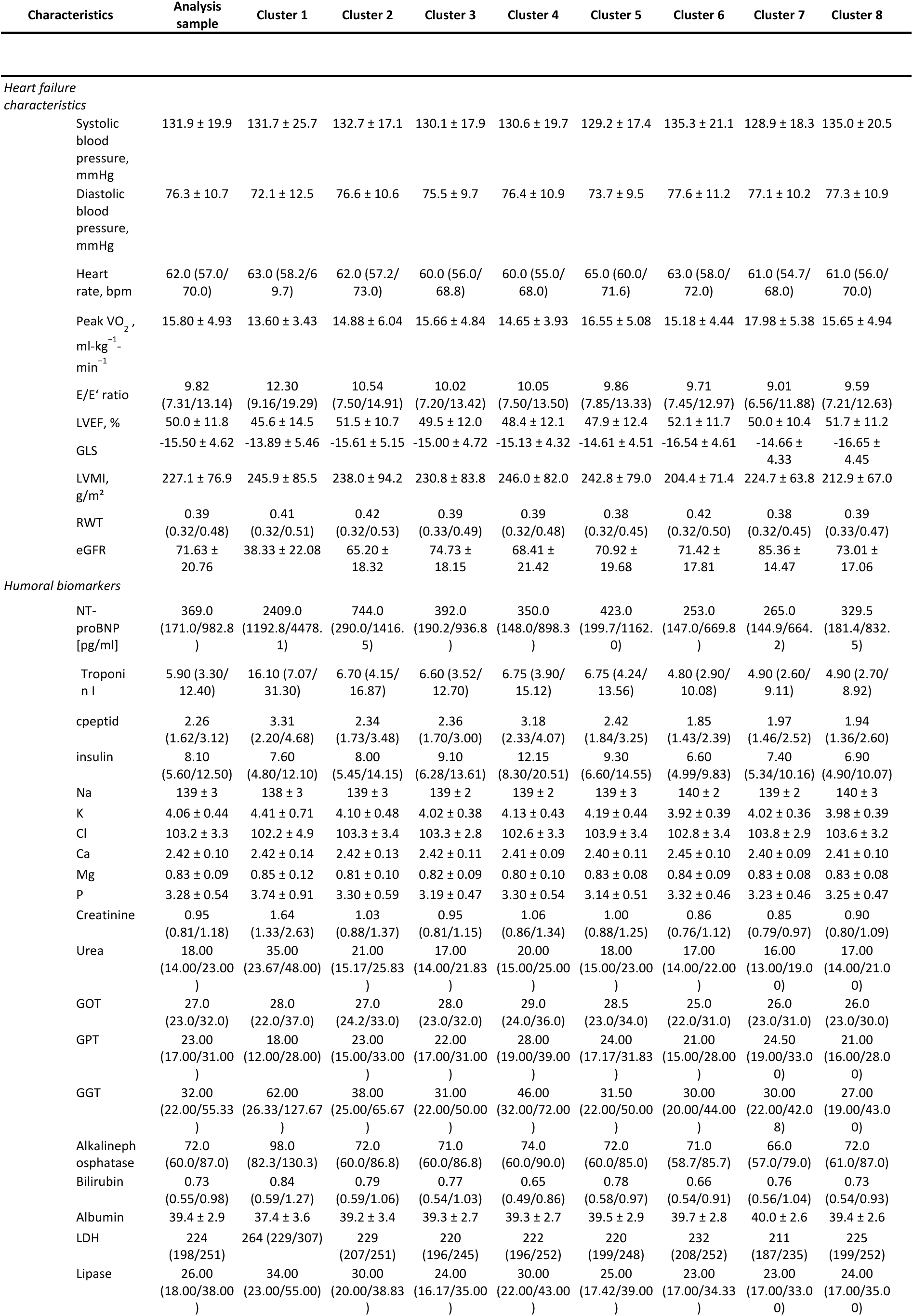
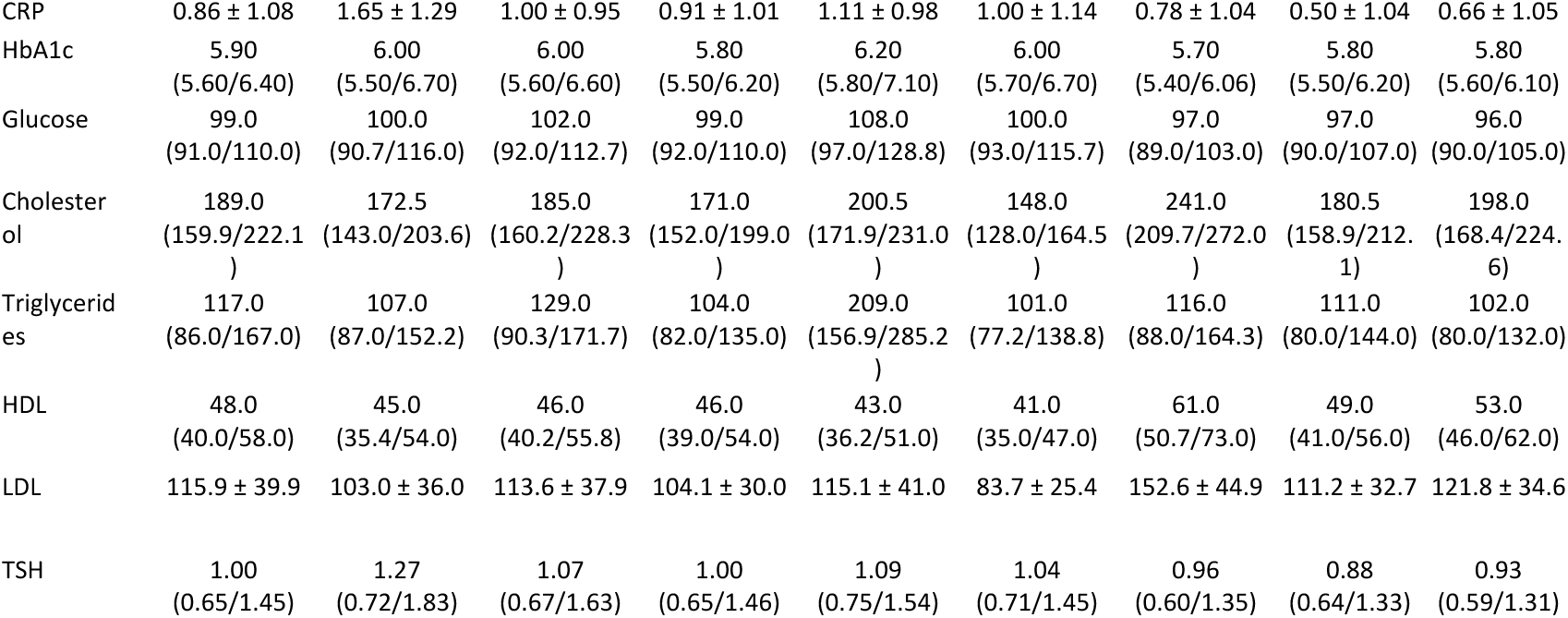
Baseline characteristics of clustering-based subgroups (continuous variables)

**Supplementary Table 3.** Cell count mean values of clustering-based subgroups.

| Cell type | Analysis sample | Cluster 1 | Cluster 2 | Cluster 3 | Cluster 4 | Cluster 5 | Cluster 6 | Cluster 7 | Cluster 8 |
| --- | --- | --- | --- | --- | --- | --- | --- | --- | --- |
| Erythrocytes | 4.55 +/- 0.48 | 4.18 +/- 0.69 | 4.51 +/- 0.63 | 4.59 +/- 0.42 | 4.55 +/- 0.48 | 4.62 +/- 0.42 | 4.48 +/- 0.44 | 4.66 +/- 0.42 | 4.57 +/- 0.47 |
| Leukocytes | 7.30<br>(6.21/8.71) | 7.46<br>(6.09/9.04) | 7.81<br>(6.65/9.15) | 7.57<br>(6.48/9.09) | 7.66<br>(6.51/9.16) | 7.57<br>(6.36/8.75) | 6.84<br>(6.03/8.11) | 7.37<br>(6.21/8.67) | 6.89<br>(5.92/8.30) |
| Hemoglobin | 13.99 +/- 1.44 | 12.72 +/- 1.75 | 13.75 +/- 1.79 | 14.19 +/- 1.40 | 14.14 +/- 1.48 | 14.18 +/- 1.35 | 13.80 +/- 1.27 | 14.34 +/- 1.20 | 13.91 +/- 1.35 |
| Hematocrit | 41.7 +/- 4.0 | 38.7 +/- 5.2 | 41.2 +/- 4.9 | 42.2 +/- 3.9 | 41.8 +/- 4.0 | 42.4 +/- 3.8 | 41.2 +/- 3.5 | 42.6 +/- 3.3 | 41.6 +/- 3.8 |
| MCV | 91.9 +/- 5.6 | 93.2 +/- 7.3 | 91.9 +/- 6.3 | 92.0 +/- 6.0 | 92.2 +/- 6.2 | 92.1 +/- 6.5 | 92.2 +/- 4.5 | 91.6 +/- 4.3 | 91.3 +/- 5.0 |
| MCH | 30.8 +/- 2.1 | 30.7 +/- 2.8 | 30.6 +/- 2.4 | 30.9 +/- 2.2 | 31.2 +/- 2.4 | 30.8 +/- 2.4 | 30.9 +/- 1.8 | 30.9 +/- 1.7 | 30.5 +/- 1.9 |
| MCHC | 33.5 +/- 1.0 | 32.9 +/- 1.3 | 33.3 +/- 1.2 | 33.6 +/- 1.0 | 33.8 +/- 1.1 or | 33.4 +/- 1.0 | 33.5 +/- 1.0 | 33.7 +/- 1.0 | 33.4 +/- 1.0 |
| EVB (red blood cell distribution) | 13.7<br>(13.2/14.4) | 14.8<br>(13.9/16.2) | 13.9<br>(13.2/14.7) | 13.7<br>(13.2/14.5) | 13.8<br>(13.2/14.5) | 13.7<br>(13.2/14.7) | 13.6<br>(13.1/14.2) | 13.5<br>(13.1/14.1) | 13.6<br>(13.2/14.3) |
| Thrombocytes | 223.6 +/- 63.8 | 206.2 +/- 78.0 | 223.7 +/- 66.4 | 227.4 +/- 62.6 | 227.1 +/- 65.9 | 207.2 +/- 60.4 | 241.3 +/- 62.0 | 215.5 +/- 59.7 | 225.1 +/- 60.7 |
| MTV | 7.72 +/- 0.93 | 7.99 +/- 0.96 | 7.82 +/- 1.09 | 7.64 +/- 1.01 | 7.62 +/- 0.91 | 7.82 +/- 0.97 | 7.66 +/- 0.88 | 7.77 +/- 0.95 | 7.72 +/- 0.84 |
| Neutrophils | 65.4 +/- 8.7 | 69.0 +/- 10.6 | 65.8 +/- 8.2 | 65.3 +/- 8.5 | 63.4 +/- 8.6 | 66.5 +/- 8.0 | 65.1 +/- 8.7 | 65.2 +/- 8.3 | 65.8 +/- 8.6 |
| Lymphocytes | 23.10<br>(18.22/28.20) | 17.65<br>(12.82/23.96) | 24.20<br>(17.30/27.35) | 23.30<br>(18.42/27.58) | 24.20<br>(19.33/29.60) | 21.40<br>(17.18/26.38) | 23.80<br>(18.97/29.20) | 23.40<br>(19.00/28.72) | 23.10<br>(18.14/28.30) |
| Monocytes | 6.00<br>(5.10/7.20) | 6.35<br>(5.40/7.66) | 6.30<br>(5.43/7.30) | 6.20<br>(5.10/7.20) | 6.20<br>(5.40/7.20) | 6.50<br>(5.40/7.58) | 5.70<br>(4.90/6.80) | 6.00<br>(5.00/7.00) | 5.90<br>(4.94/7.10) |
| Eosinophils | 2.10<br>(1.30/3.10) | 2.50<br>(1.50/4.08) | 1.90<br>(1.20/2.73) | 2.05<br>(1.29/3.00) | 2.25<br>(1.40/3.30) | 2.00<br>(1.29/3.00) | 2.10<br>(1.34/3.40) | 2.00<br>(1.20/2.70) | 2.00<br>(1.30/2.90) |
| Basophils | 0.50<br>(0.40/0.70) | 0.60<br>(0.40/0.78) | 0.55<br>(0.40/0.70) | 0.60<br>(0.40/0.78) | 0.60<br>(0.40/0.80) | 0.50<br>(0.40/0.70) | 0.50<br>(0.40/0.70) | 0.50<br>(0.40/0.70) | 0.50<br>(0.40/0.70) |

**Supplementary Table 4.**
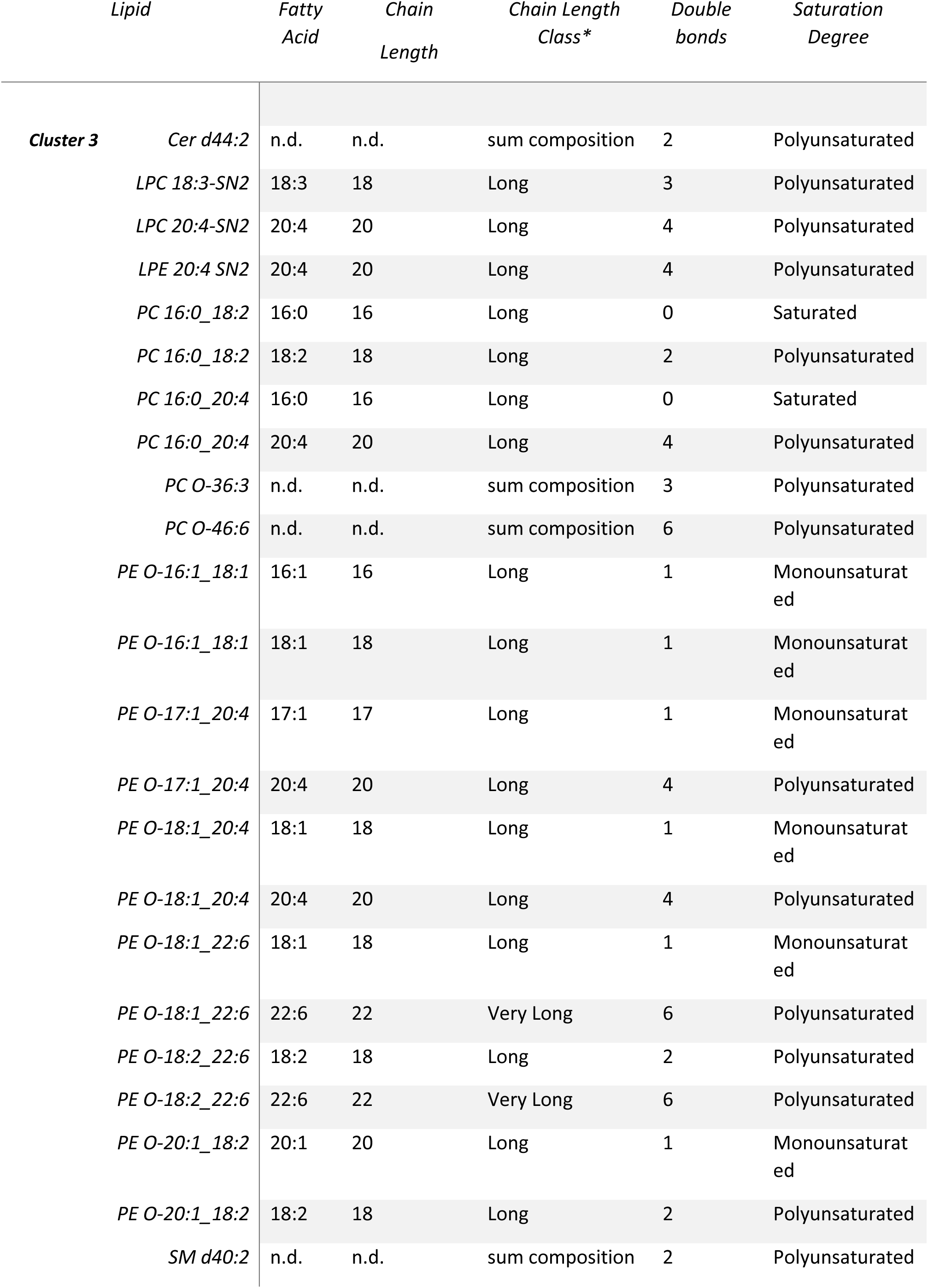
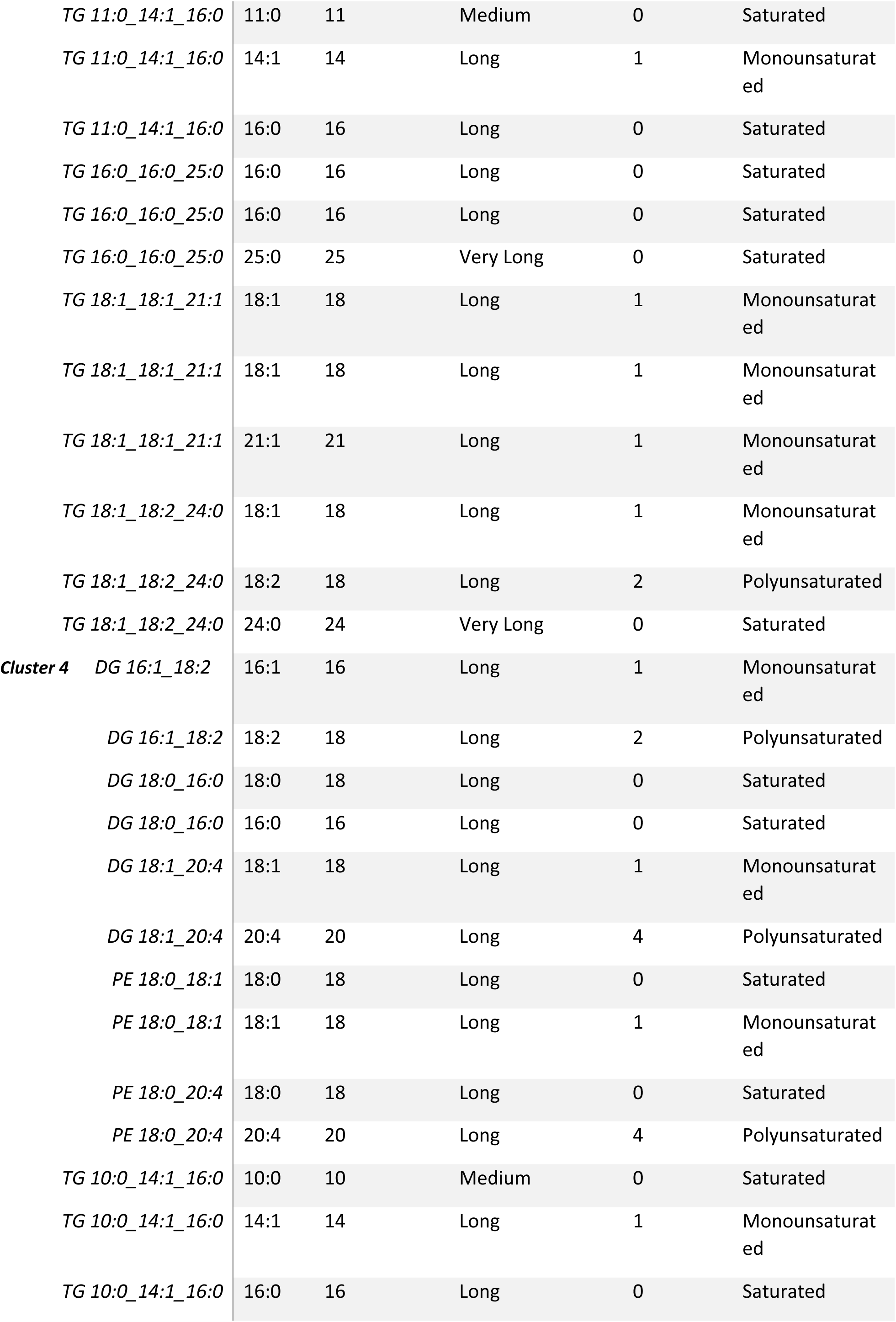
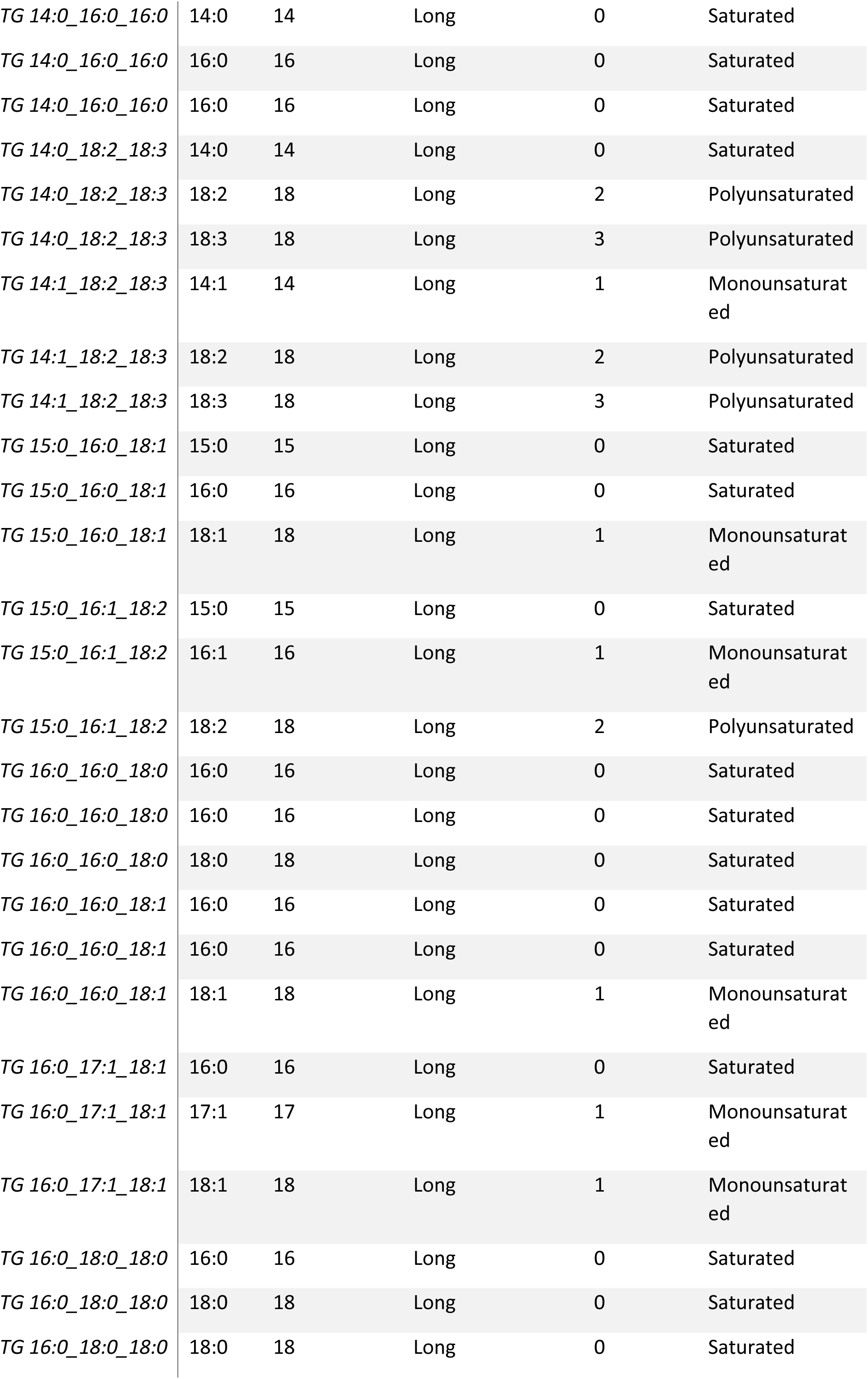
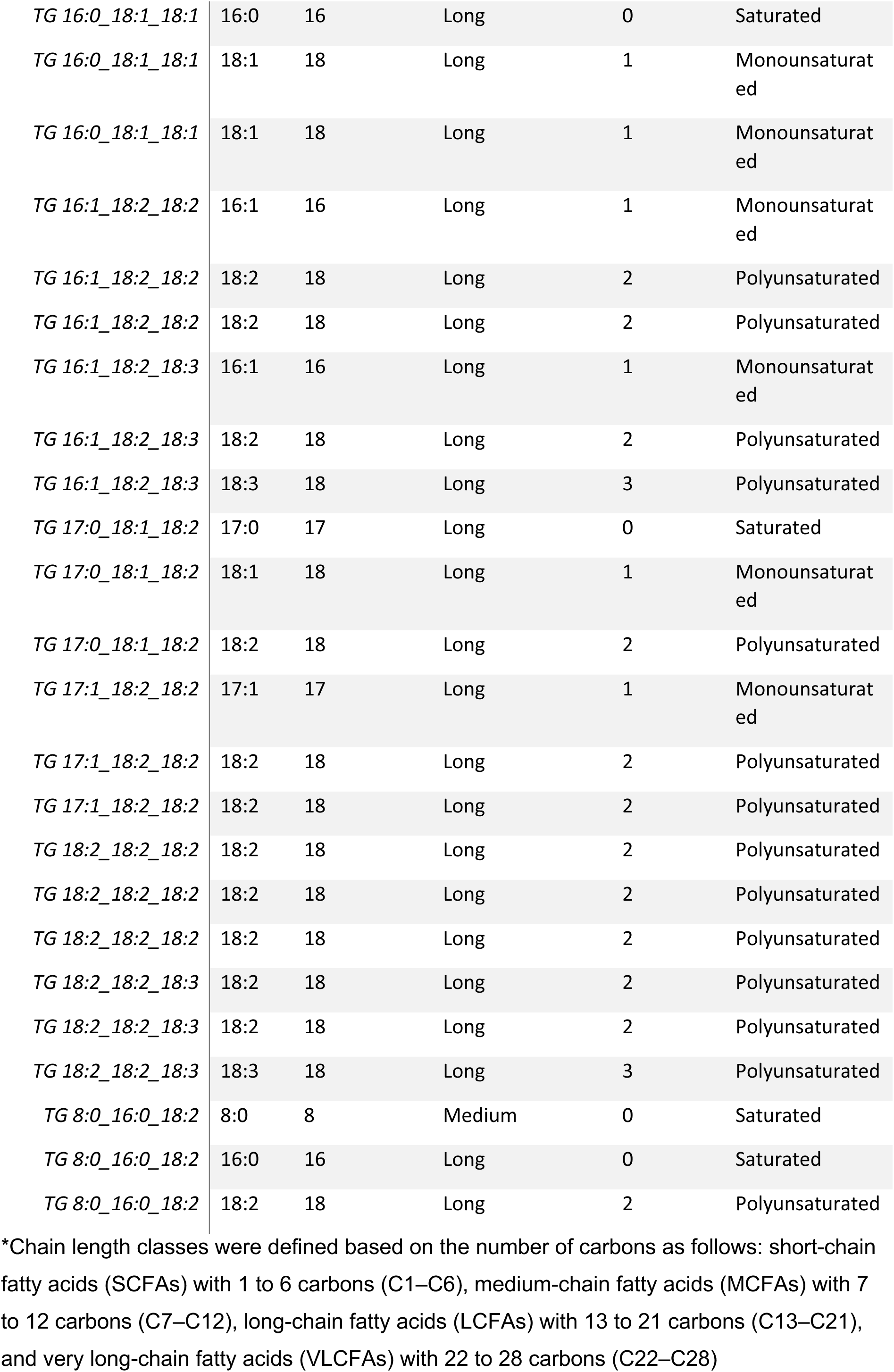
Fatty acyls chain length and saturation degree.

| Lipid |  | Fatty Acid | Chain Length | Chain Length Class* | Double bonds | Saturation Degree |
| --- | --- | --- | --- | --- | --- | --- |
| Cluster 3 | Cer d44:2 | n.d. | n.d. | sum composition | 2 | Polyunsaturated |
|  | LPC 18:3-SN2 | 18:3 | 18 | Long | 3 | Polyunsaturated |
|  | LPC 20:4-SN2 | 20:4 | 20 | Long | 4 | Polyunsaturated |
|  | LPE 20:4 SN2 | 20:4 | 20 | Long | 4 | Polyunsaturated |
|  | PC 16:0_18:2 | 16:0 | 16 | Long | 0 | Saturated |
|  | PC 16:0_18:2 | 18:2 | 18 | Long | 2 | Polyunsaturated |
|  | PC 16:0_20:4 | 16:0 | 16 | Long | 0 | Saturated |
|  | PC 16:0_20:4 | 20:4 | 20 | Long | 4 | Polyunsaturated |
|  | PC O-36:3 | n.d. | n.d. | sum composition | 3 | Polyunsaturated |
|  | PC O-46:6 | n.d. | n.d. | sum composition | 6 | Polyunsaturated |
|  | PE O-16:1_18:1 | 16:1 | 16 | Long | 1 | Monounsaturated |
|  | PE O-16:1_18:1 | 18:1 | 18 | Long | 1 | Monounsaturated |
|  | PE O-17:1_20:4 | 17:1 | 17 | Long | 1 | Monounsaturated |
|  | PE O-17:1_20:4 | 20:4 | 20 | Long | 4 | Polyunsaturated |
|  | PE O-18:1_20:4 | 18:1 | 18 | Long | 1 | Monounsaturated |
|  | PE O-18:1_20:4 | 20:4 | 20 | Long | 4 | Polyunsaturated |
|  | PE O-18:1_22:6 | 18:1 | 18 | Long | 1 | Monounsaturated |
|  | PE O-18:1_22:6 | 22:6 | 22 | Very Long | 6 | Polyunsaturated |
|  | PE O-18:2_22:6 | 18:2 | 18 | Long | 2 | Polyunsaturated |
|  | PE O-18:2_22:6 | 22:6 | 22 | Very Long | 6 | Polyunsaturated |
|  | PE O-20:1_18:2 | 20:1 | 20 | Long | 1 | Monounsaturated |
|  | PE O-20:1_18:2 | 18:2 | 18 | Long | 2 | Polyunsaturated |
|  | SM d40:2 | n.d. | n.d. | sum composition | 2 | Polyunsaturated |

|  |  |  |  |  |  |
| --- | --- | --- | --- | --- | --- |
| *TG 11:0_14:1_16:0* | 11:0 | 11 | Medium | 0 | Saturated |
| *TG 11:0_14:1_16:0* | 14:1 | 14 | Long | 1 | Monounsaturated |
| *TG 11:0_14:1_16:0* | 16:0 | 16 | Long | 0 | Saturated |
| *TG 16:0_16:0_25:0* | 16:0 | 16 | Long | 0 | Saturated |
| *TG 16:0_16:0_25:0* | 16:0 | 16 | Long | 0 | Saturated |
| *TG 16:0_16:0_25:0* | 25:0 | 25 | Very Long | 0 | Saturated |
| *TG 18:1_18:1_21:1* | 18:1 | 18 | Long | 1 | Monounsaturated |
| *TG 18:1_18:1_21:1* | 18:1 | 18 | Long | 1 | Monounsaturated |
| *TG 18:1_18:1_21:1* | 21:1 | 21 | Long | 1 | Monounsaturated |
| *TG 18:1_18:2_24:0* | 18:1 | 18 | Long | 1 | Monounsaturated |
| *TG 18:1_18:2_24:0* | 18:2 | 18 | Long | 2 | Polyunsaturated |
| *TG 18:1_18:2_24:0* | 24:0 | 24 | Very Long | 0 | Saturated |
| **Cluster 4** *DG 16:1_18:2* | 16:1 | 16 | Long | 1 | Monounsaturated |
| *DG 16:1_18:2* | 18:2 | 18 | Long | 2 | Polyunsaturated |
| *DG 18:0_16:0* | 18:0 | 18 | Long | 0 | Saturated |
| *DG 18:0_16:0* | 16:0 | 16 | Long | 0 | Saturated |
| *DG 18:1_20:4* | 18:1 | 18 | Long | 1 | Monounsaturated |
| *DG 18:1_20:4* | 20:4 | 20 | Long | 4 | Polyunsaturated |
| *PE 18:0_18:1* | 18:0 | 18 | Long | 0 | Saturated |
| *PE 18:0_18:1* | 18:1 | 18 | Long | 1 | Monounsaturated |
| *PE 18:0_20:4* | 18:0 | 18 | Long | 0 | Saturated |
| *PE 18:0_20:4* | 20:4 | 20 | Long | 4 | Polyunsaturated |
| *TG 10:0_14:1_16:0* | 10:0 | 10 | Medium | 0 | Saturated |
| *TG 10:0_14:1_16:0* | 14:1 | 14 | Long | 1 | Monounsaturated |
| *TG 10:0_14:1_16:0* | 16:0 | 16 | Long | 0 | Saturated |
| *TG 14:0_16:0_16:0* | 14:0 | 14 | Long | 0 | Saturated |
| *TG 14:0_16:0_16:0* | 16:0 | 16 | Long | 0 | Saturated |
| *TG 14:0_16:0_16:0* | 16:0 | 16 | Long | 0 | Saturated |
| *TG 14:0_18:2_18:3* | 14:0 | 14 | Long | 0 | Saturated |
| *TG 14:0_18:2_18:3* | 18:2 | 18 | Long | 2 | Polyunsaturated |
| *TG 14:0_18:2_18:3* | 18:3 | 18 | Long | 3 | Polyunsaturated |
| *TG 14:1_18:2_18:3* | 14:1 | 14 | Long | 1 | Monounsaturated |
| *TG 14:1_18:2_18:3* | 18:2 | 18 | Long | 2 | Polyunsaturated |
| *TG 14:1_18:2_18:3* | 18:3 | 18 | Long | 3 | Polyunsaturated |
| *TG 15:0_16:0_18:1* | 15:0 | 15 | Long | 0 | Saturated |
| *TG 15:0_16:0_18:1* | 16:0 | 16 | Long | 0 | Saturated |
| *TG 15:0_16:0_18:1* | 18:1 | 18 | Long | 1 | Monounsaturated |
| *TG 15:0_16:1_18:2* | 15:0 | 15 | Long | 0 | Saturated |
| *TG 15:0_16:1_18:2* | 16:1 | 16 | Long | 1 | Monounsaturated |
| *TG 15:0_16:1_18:2* | 18:2 | 18 | Long | 2 | Polyunsaturated |
| *TG 16:0_16:0_18:0* | 16:0 | 16 | Long | 0 | Saturated |
| *TG 16:0_16:0_18:0* | 16:0 | 16 | Long | 0 | Saturated |
| *TG 16:0_16:0_18:0* | 18:0 | 18 | Long | 0 | Saturated |
| *TG 16:0_16:0_18:1* | 16:0 | 16 | Long | 0 | Saturated |
| *TG 16:0_16:0_18:1* | 16:0 | 16 | Long | 0 | Saturated |
| *TG 16:0_16:0_18:1* | 18:1 | 18 | Long | 1 | Monounsaturated |
| *TG 16:0_17:1_18:1* | 16:0 | 16 | Long | 0 | Saturated |
| *TG 16:0_17:1_18:1* | 17:1 | 17 | Long | 1 | Monounsaturated |
| *TG 16:0_17:1_18:1* | 18:1 | 18 | Long | 1 | Monounsaturated |
| *TG 16:0_18:0_18:0* | 16:0 | 16 | Long | 0 | Saturated |
| *TG 16:0_18:0_18:0* | 18:0 | 18 | Long | 0 | Saturated |
| *TG 16:0_18:0_18:0* | 18:0 | 18 | Long | 0 | Saturated |
| *TG 16:0_18:1_18:1* | 16:0 | 16 | Long | 0 | Saturated |
| *TG 16:0_18:1_18:1* | 18:1 | 18 | Long | 1 | Monounsaturated |
| *TG 16:0_18:1_18:1* | 18:1 | 18 | Long | 1 | Monounsaturated |
| *TG 16:1_18:2_18:2* | 16:1 | 16 | Long | 1 | Monounsaturated |
| *TG 16:1_18:2_18:2* | 18:2 | 18 | Long | 2 | Polyunsaturated |
| *TG 16:1_18:2_18:2* | 18:2 | 18 | Long | 2 | Polyunsaturated |
| *TG 16:1_18:2_18:3* | 16:1 | 16 | Long | 1 | Monounsaturated |
| *TG 16:1_18:2_18:3* | 18:2 | 18 | Long | 2 | Polyunsaturated |
| *TG 16:1_18:2_18:3* | 18:3 | 18 | Long | 3 | Polyunsaturated |
| *TG 17:0_18:1_18:2* | 17:0 | 17 | Long | 0 | Saturated |
| *TG 17:0_18:1_18:2* | 18:1 | 18 | Long | 1 | Monounsaturated |
| *TG 17:0_18:1_18:2* | 18:2 | 18 | Long | 2 | Polyunsaturated |
| *TG 17:1_18:2_18:2* | 17:1 | 17 | Long | 1 | Monounsaturated |
| *TG 17:1_18:2_18:2* | 18:2 | 18 | Long | 2 | Polyunsaturated |
| *TG 17:1_18:2_18:2* | 18:2 | 18 | Long | 2 | Polyunsaturated |
| *TG 18:2_18:2_18:2* | 18:2 | 18 | Long | 2 | Polyunsaturated |
| *TG 18:2_18:2_18:2* | 18:2 | 18 | Long | 2 | Polyunsaturated |
| *TG 18:2_18:2_18:2* | 18:2 | 18 | Long | 2 | Polyunsaturated |
| *TG 18:2_18:2_18:3* | 18:2 | 18 | Long | 2 | Polyunsaturated |
| *TG 18:2_18:2_18:3* | 18:2 | 18 | Long | 2 | Polyunsaturated |
| *TG 18:2_18:2_18:3* | 18:3 | 18 | Long | 3 | Polyunsaturated |
| *TG 8:0_16:0_18:2* | 8:0 | 8 | Medium | 0 | Saturated |
| *TG 8:0_16:0_18:2* | 16:0 | 16 | Long | 0 | Saturated |
| *TG 8:0_16:0_18:2* | 18:2 | 18 | Long | 2 | Polyunsaturated |

\*Chain length classes were defined based on the number of carbons as follows: short-chain fatty acids (SCFAs) with 1 to 6 carbons (C1–C6), medium-chain fatty acids (MCFAs) with 7 to 12 carbons (C7–C12), long-chain fatty acids (LCFAs) with 13 to 21 carbons (C13–C21), and very long-chain fatty acids (VLCFAs) with 22 to 28 carbons (C22–C28)

**Supplementary Figure 1.**
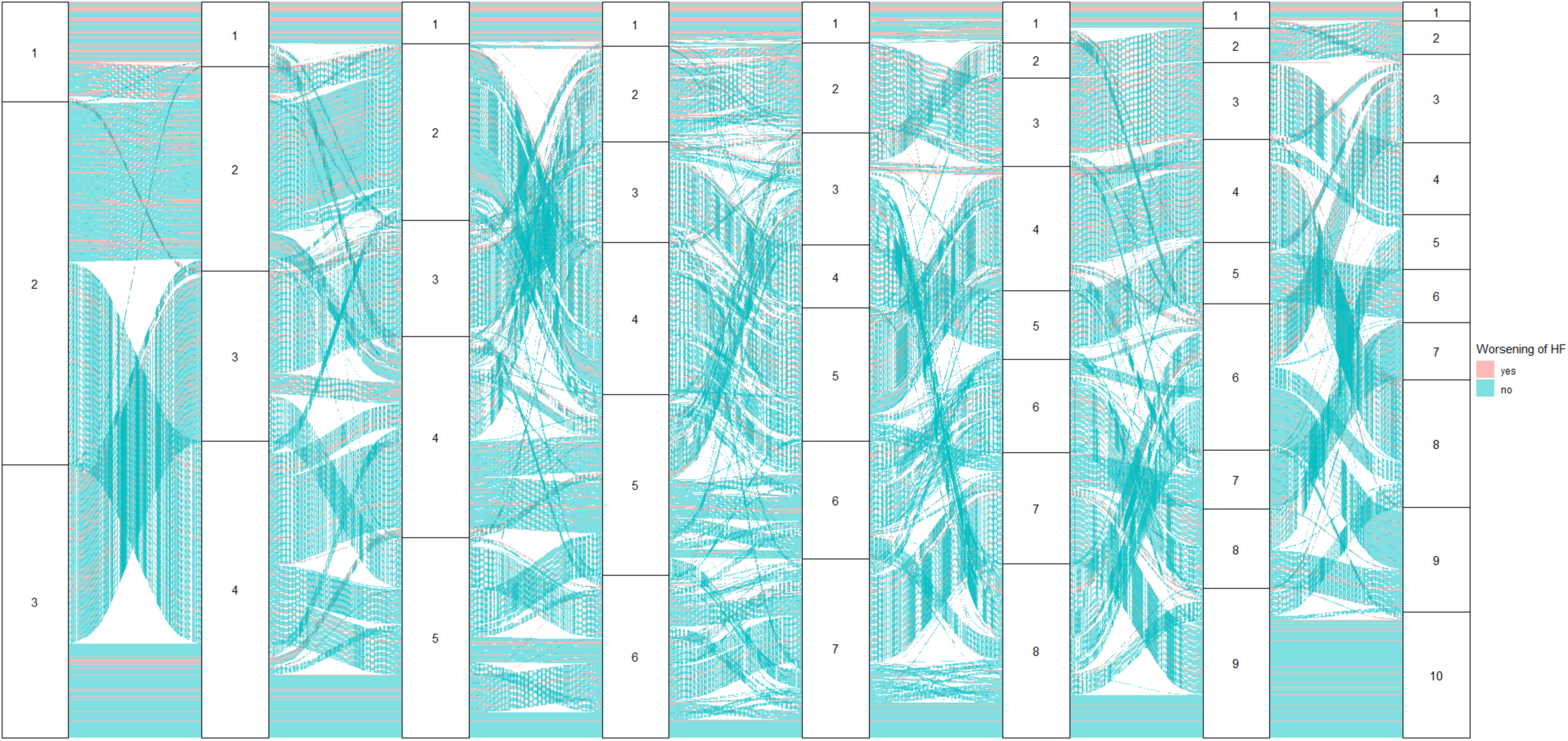
Sankey plot of 1,678 subjects with C or D stage heart failure, clustered from 3 to 10 clusters. The plot shows the flow and changes in combination of subjects in each cluster, based on the chosen number of clusters. The color is coded for worsening of heart failure.

**Supplementary Figure 2.**
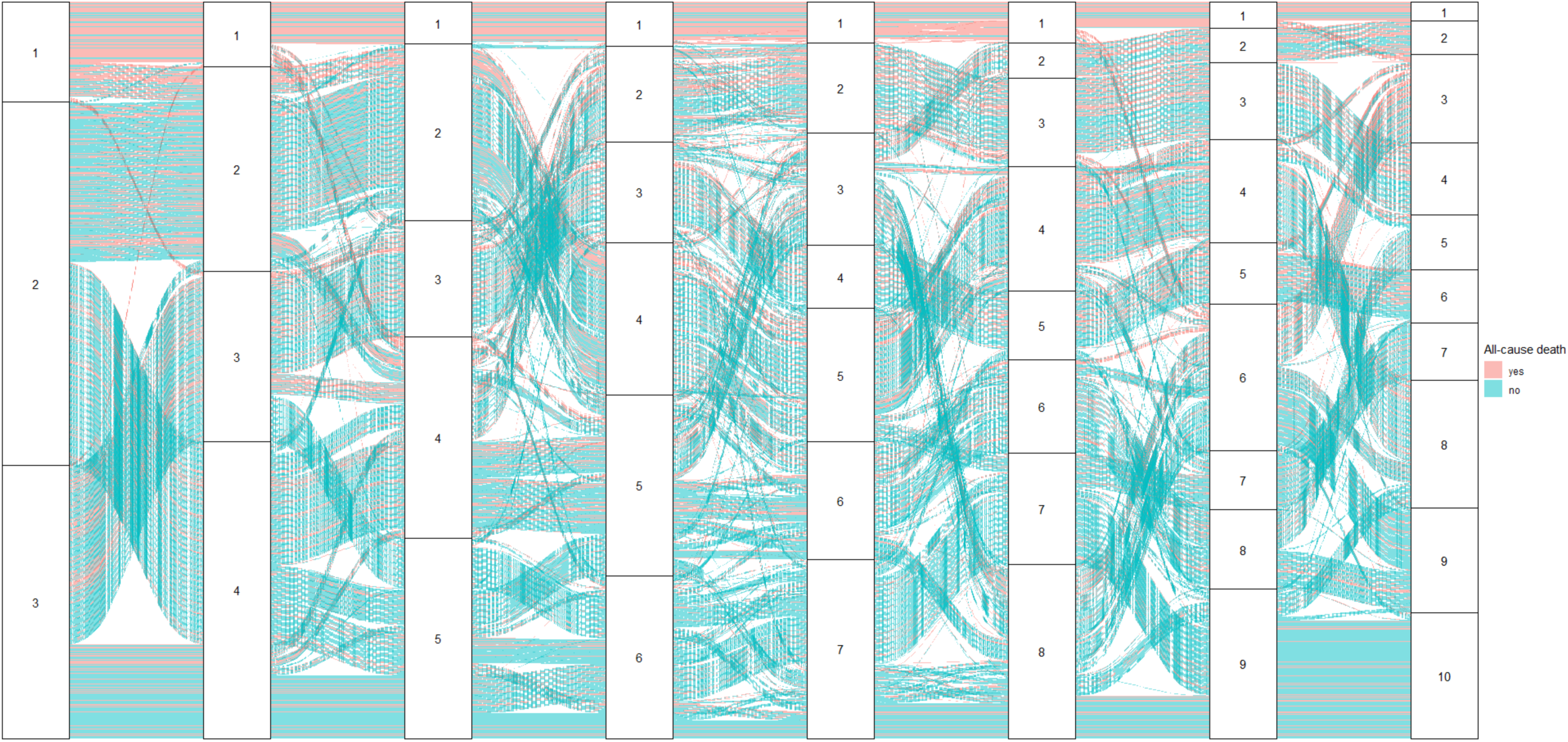
Sankey plot of 1,678 subjects with C or D stage heart failure, clustered from 3 to 10 clusters. The plot shows the flow and changes in combination of subjects in each cluster, based on the chosen number of clusters. The color is coded for all-cause death.

**Supplementary Figure 3.**
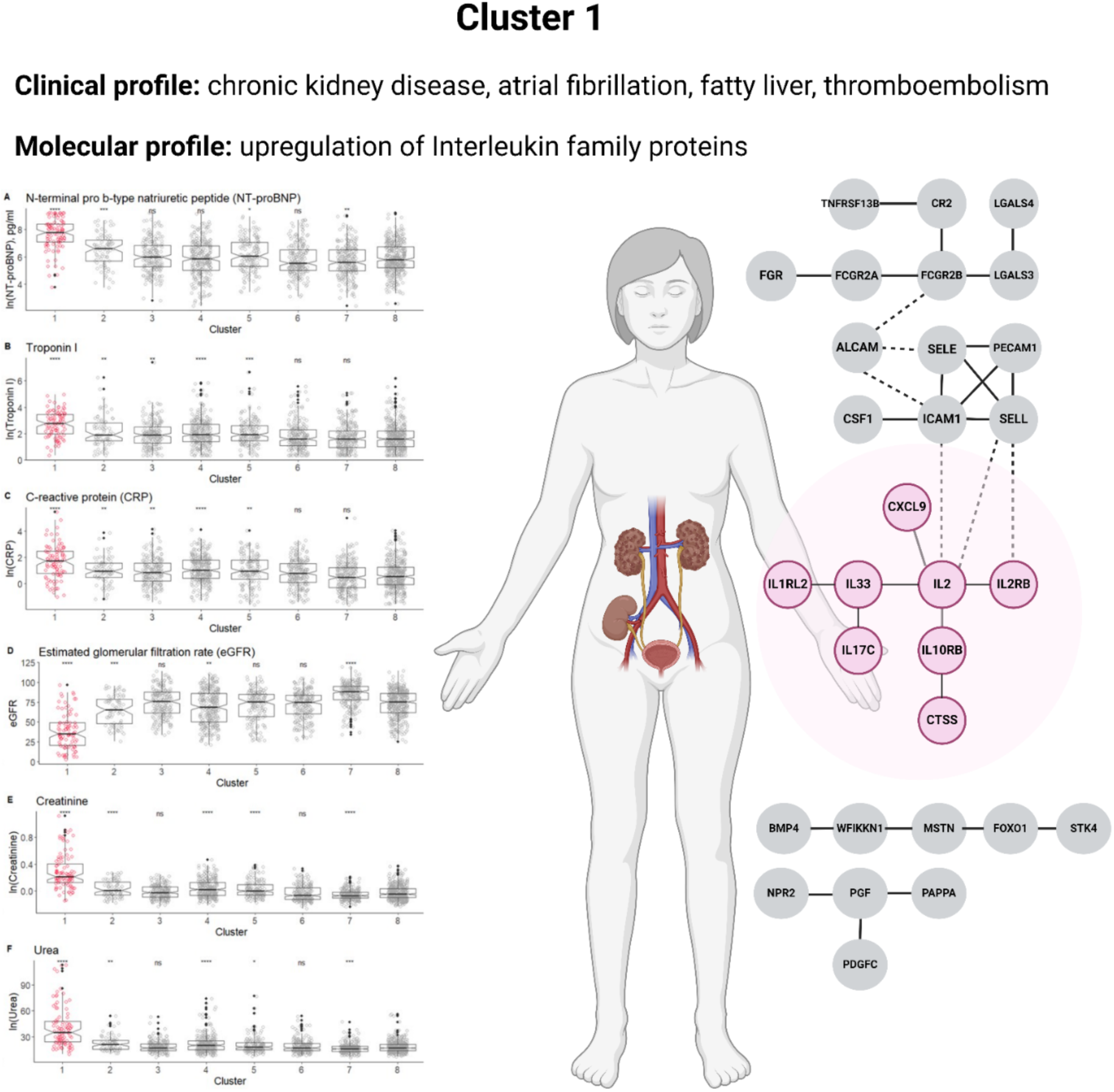
Molecular and clinical characterization of the worse outcome clusters - Cluster 1.

**Supplementary Figure 4.**
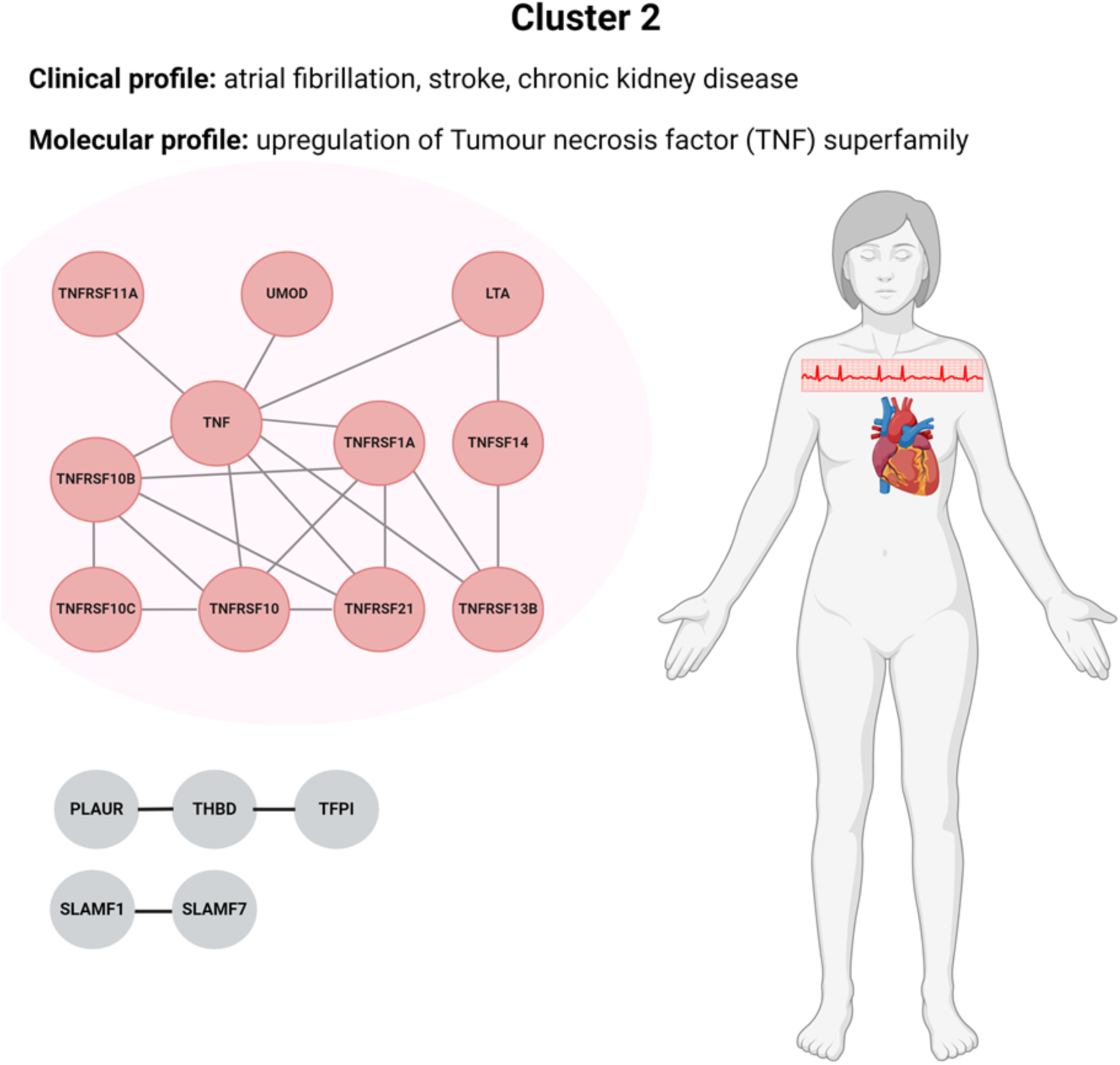
Molecular and clinical characterization of the worse outcome clusters - Cluster 2.

**Supplementary Figure 5.**
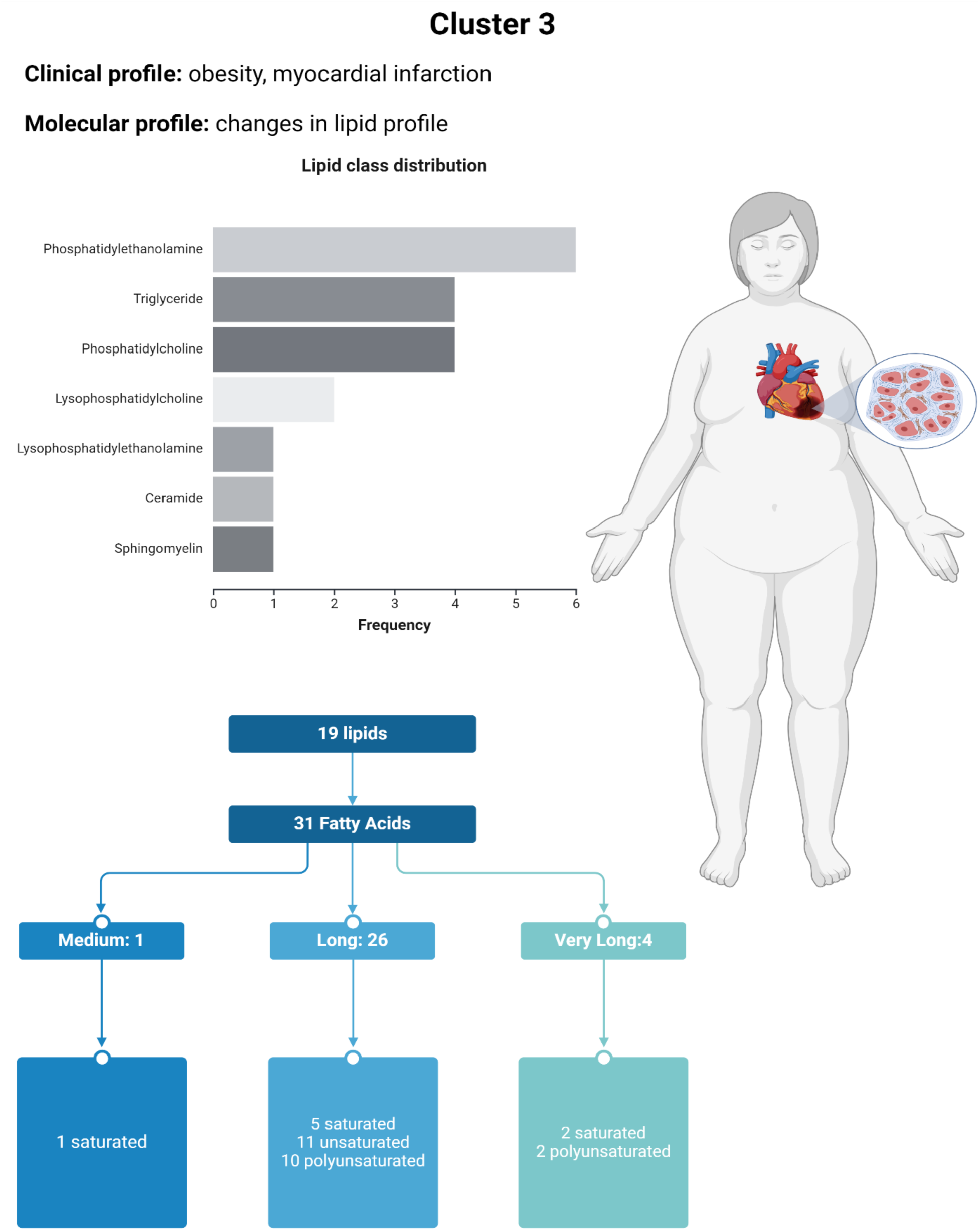
Molecular and clinical characterization of the worse outcome clusters - Cluster 3. Chain length classes were defined based on the number of carbons as follows: short-chain fatty acids (SCFAs) with 1 to 6 carbons (C1–C6), medium-chain fatty acids (MCFAs) with 7 to 12 carbons (C7–C12), long-chain fatty acids (LCFAs) with 13 to 21 carbons (C13–C21), and very long-chain fatty acids (VLCFAs) with 22 to 28 carbons (C22–C28)

**Supplementary Figure 6.**
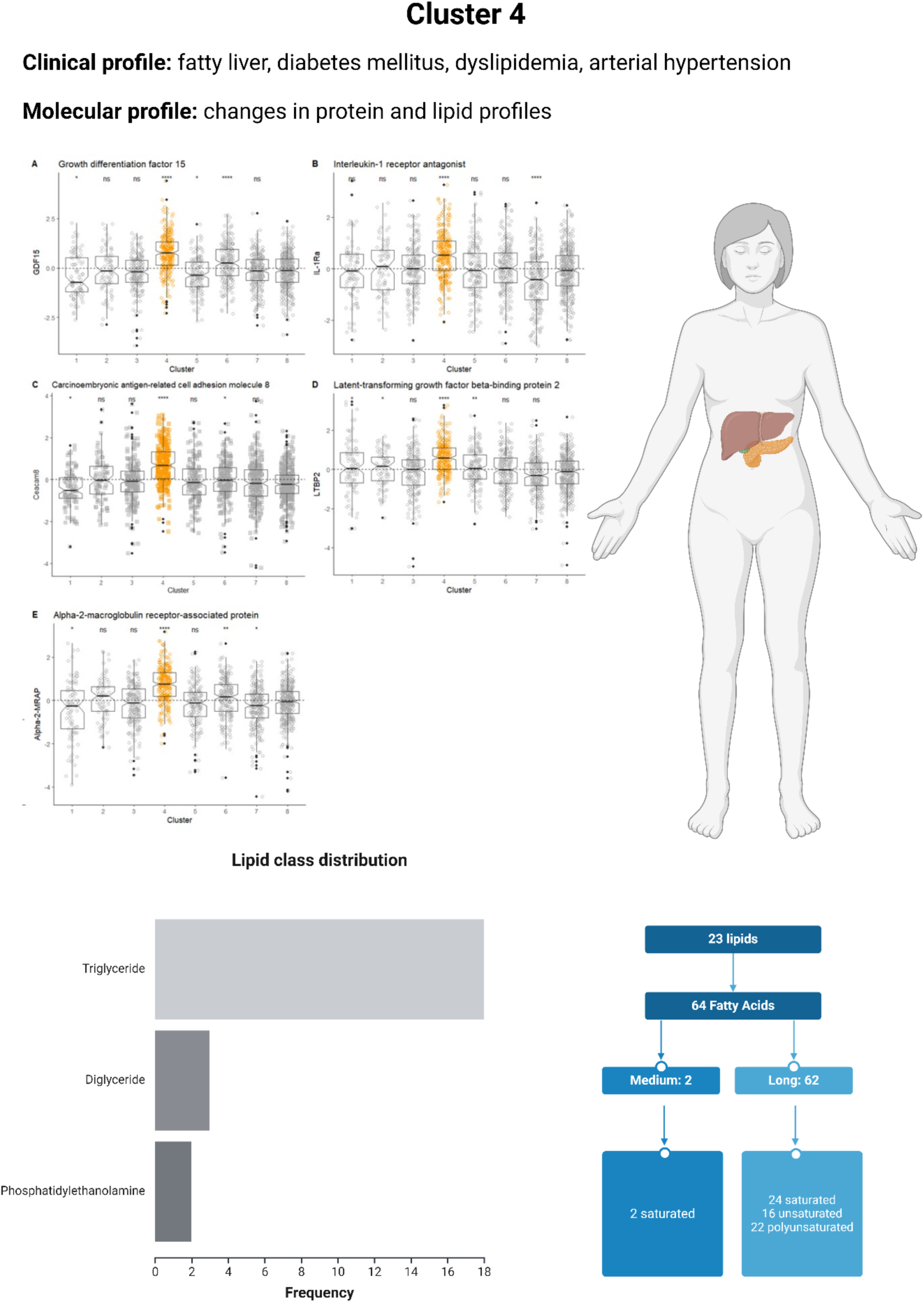
Molecular and clinical characterization of the worse outcome clusters - Cluster 4. Chain length classes were defined based on the number of carbons as follows: short-chain fatty acids (SCFAs) with 1 to 6 carbons (C1–C6), medium-chain fatty acids (MCFAs) with 7 to 12 carbons (C7–C12), long-chain fatty acids (LCFAs) with 13 to

**Supplementary Figure 7.**
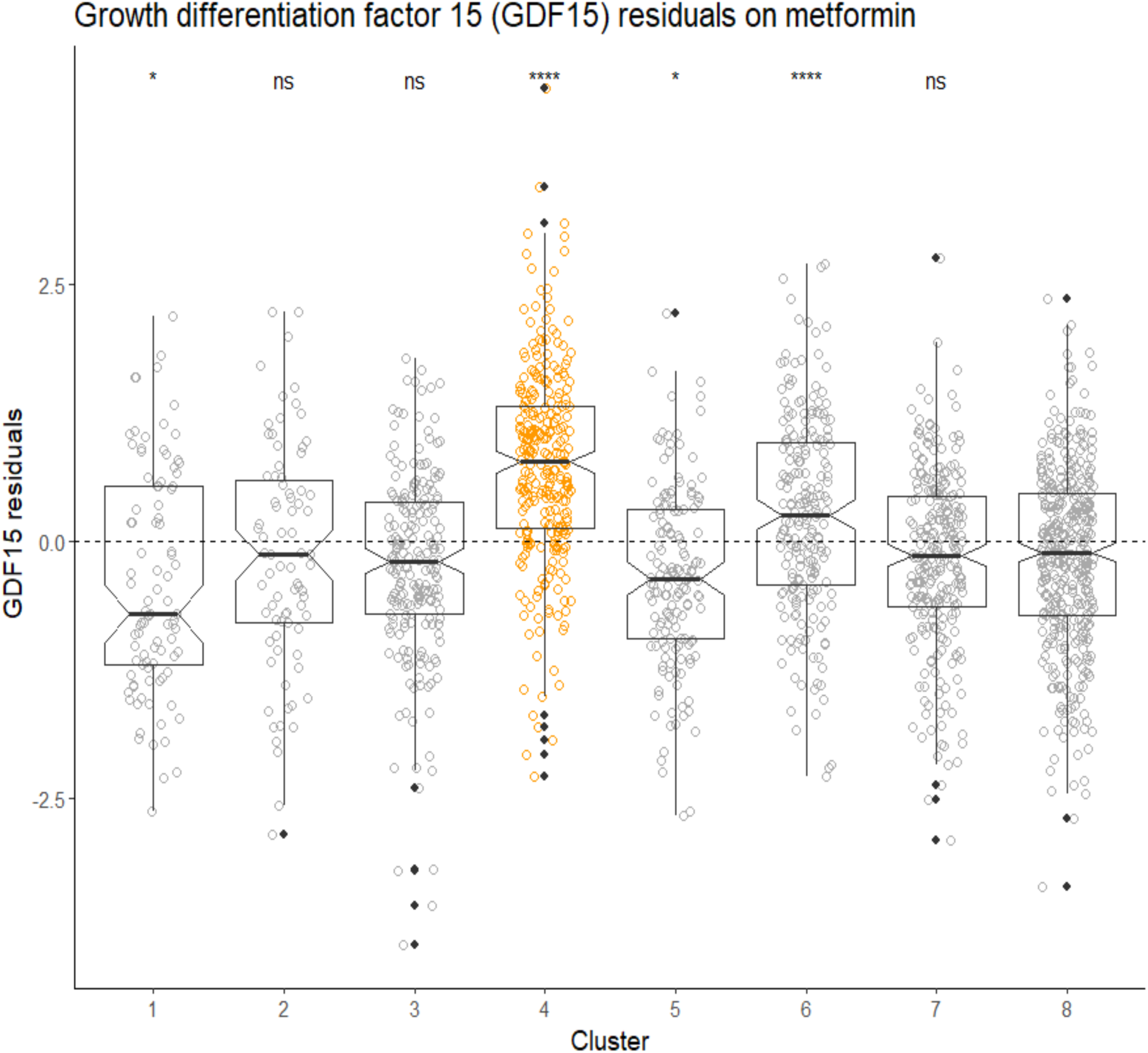
Residuals from the regression of GDF-15 expression on metformin intake (binary variable). The residuals still demonstrated the significant increase of GDF-15 in cluster 4, pointing at the additional independent of metformin intake mechanisms of this upregulation.

